# Prognostic value of early leukocyte fluctuations for recovery from traumatic spinal cord injury

**DOI:** 10.1101/2020.10.26.20220236

**Authors:** T Jogia, T Lübstorf, E Jacobson, E Scriven, S Atresh, T Liebscher, JM Schwab, MA Kopp, J Walsham, KE Campbell, MJ Ruitenberg

## Abstract

**Background:** Acute traumatic spinal cord injury (SCI) induces a systemic immune response involving circulating white blood cells (WBC). How this response is influenced by overall trauma severity, the neurological level of injury and/or correlates with patient outcomes is poorly understood. The objective of this study was to identify relationships between early changes in circulating WBCs, injury characteristics, and long-term patient outcomes in individuals with traumatic SCI.

**Methods:** We retrospectively analysed data from n=161 SCI patients admitted to Brisbane’s Princess Alexandra Hospital (exploration cohort). Logistic regression models in conjunction with receiver-operator characteristic (ROC) analyses were used to assess the strength of specific links between the WBC response, respiratory infection incidence and neurological outcomes (American Spinal Injury Association Impairment Scale (AIS) grade conversion). An independent validation cohort from the Trauma Hospital Berlin, Germany (n=49) was then probed to assess the robustness of effects and to disentangle centre effects.

**Results:** We find that the extent of acute neutrophilia in human SCI patients is positively correlated with New Injury Severity Scores (NISS) but inversely with the neurological outcome (AIS grade). Multivariate analysis demonstrated that acute SCI-induced neutrophilia is an independent predictor of AIS grade conversion failure, with an odds ratio (OR) of 4.16 and ROC area under curve (AUC) of 0.82 (p<0.0001). SCI-induced lymphopenia was separately identified as an independent predictor of better recovery (OR = 24.15; ROC AUC = 0.85, p<0.0001). Acute neutrophilia and increased neutrophil-lymphocyte ratios were otherwise significantly associated with respiratory infection presentation in both patient cohorts.

**Conclusions:** Our findings demonstrate the prognostic value of modelling early circulating neutrophil and lymphocyte counts with patient characteristics for predicting the longer-term recovery after SCI.

## INTRODUCTION

Traumatic spinal cord injury (SCI) is hallmarked by a chronic, non-resolving inflammatory response at the lesion site that involves both CNS-resident and infiltrating white blood cells.^1,2^ Although reparative aspects have been ascribed to at least some aspects of inflammation after SCI,^3-5^ on balance, the inflammatory response is thought to be maladaptive and an amplifier of secondary pathology in spared spinal cord tissue.^6^ Numerous experimental animal studies, mostly conducted in rodents, have provided a detailed description of the spatiotemporal development of the cellular inflammatory response to SCI (reviewed in ^6^), the key features of which are recapitulated in human patients.^7^ Neutrophils and monocytes dominate in the early inflammatory infiltrate at the site of SCI, and hence these cells continue to be intensely investigated as possible targets for immunomodulation.^4,5,8,9^

Somewhat paradoxically, neural injuries affecting the brain or spinal cord are often also accompanied by a period of systemic immune depression in the periphery that can promote the development of nosocomial infections.^10,11^ In the case of SCI, this is commonly referred to as SCI-induced immune depression syndrome (SCI-IDS), a phenomenon that is conventionally linked to severe lymphopenia.^12^ It is believed that SCI-IDS increases the susceptibility of this patient population to potentially life-threatening infections, which continue to be a leading cause of death after SCI.^13^ There is also evidence to suggest that patients who acquire these infections show poorer long-term neurological recovery.^14-16^ Given that infectious complications are more prevalent in individuals with a cervical or upper thoracic SCI, it has been argued that the extent of immunosuppression is directly associated to the level of SCI, at least partially owing to disruptions in sympathetic signalling to and/or feedback signals from immune organs.^17,18^ Importantly, however, it was recently demonstrated that lymphopenia itself may not directly lead to greater infection susceptibility in experimental animal models,^17^ and the underlying mechanism driving the increased infection risk in SCI patients thus remains incompletely understood at present. A deeper understanding of how the phenomenon of SCI-IDS co-exists with and/or influences pro-inflammatory and injurious processes at the spinal cord lesion site is currently also lacking in the field.

Here we sought to document the systemic response to human traumatic SCI as part of a retrospective study in which we explored putative links between early changes in circulating WBC numbers, various concomitant injury factors such as injury level/severity,^19-21^ and longer-term neurological outcomes in an Australian cohort of SCI patients that was admitted to Brisbane’s Princess Alexandra Hospital (PAH) over a 5-year period. The prevalence of respiratory infections and their relationship to injury characteristics and systemic WBC changes were also studied in these patients to identify possible predictors of heightened infection risk and/or connections with SCI-IDS manifestation. We lastly explored whether key findings of our study could be reproduced in an independent German cohort of patients, admitted to the Trauma Hospital Berlin, in order to confirm the validity of our observations across different healthcare systems.

## METHODS

### Standard protocol approvals, registrations and patient consents

Suitable study participants were identified through the Princess Alexandra Hospital (PAH) Trauma Registry, and the relevant clinical data extracted from patient records. A total of 163 patients with a confirmed traumatic SCI, aged 15 to 98 years and admitted to PAH between 2012 and 2017, were included in this study. Demographics and baseline characteristics for this patient group (hereafter referred to as the ‘Brisbane cohort’) are detailed in *Table 1*. The retrospective analysis of SCI patient data was approved by the Queensland Government’s Human Research Ethics Committee for Metro South Health (HREC/16/QPAH/196) and ratified by The University of Queensland (HREC 2014000031). A separate cohort of 49 SCI patients over 18 years of age and admitted to the Trauma Hospital Berlin between 2011 and 2017 was used to validate key findings from the Brisbane cohort. Patient details for these individuals (hereafter referred to as the ‘Berlin cohort’) are shown in *Table 2*. To generate a comparable dataset to the Brisbane cohort, routine clinical care data of SCI patients in the Berlin cohort, as collected for the Comparative Outcome and Treatment Evaluation in SCI (COaT-SCI) study, was combined with available blood test data for the same individuals under an observational study conducted at the same centre; both studies were locally approved by the Human Research Ethics Committee of Charité-Universitätsmedizin Berlin (#EA2/015/15 and #EA1/001/09, respectively). Patients with no documented WBC data for the first week post-SCI were excluded from any further analysis in this study.

**Table 1:**
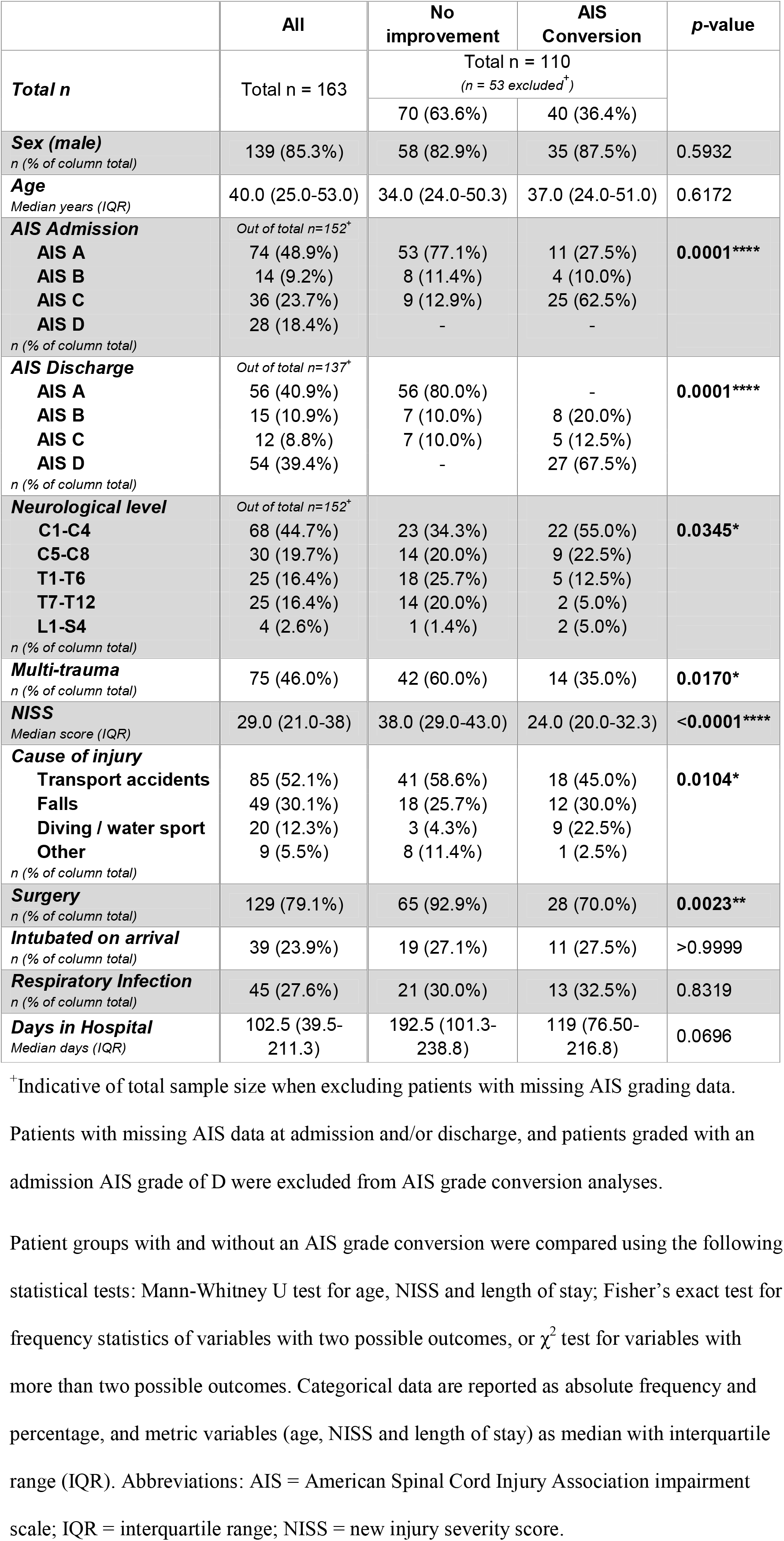
Baseline characteristics of the Brisbane patient cohort

|  | All | No improvement | AIS Conversion | p-value |
| --- | --- | --- | --- | --- |
| **Total n** | Total n = 163 | Total n = 110<br>(n = 53 excluded<sup>†</sup>) |  |  |
|  |  | 70 (63.6%) | 40 (36.4%) |  |
| **Sex (male)**<br>n (% of column total) | 139 (85.3%) | 58 (82.9%) | 35 (87.5%) | 0.5932 |
| **Age**<br>Median years (IQR) | 40.0 (25.0-53.0) | 34.0 (24.0-50.3) | 37.0 (24.0-51.0) | 0.6172 |
| **AIS Admission**<br>Out of total n=152<sup>†</sup> |  |  |  |  |
| **AIS A** | 74 (48.9%) | 53 (77.1%) | 11 (27.5%) | **0.0001\*\*\*\*** |
| **AIS B** | 14 (9.2%) | 8 (11.4%) | 4 (10.0%) |  |
| **AIS C** | 36 (23.7%) | 9 (12.9%) | 25 (62.5%) |  |
| **AIS D** | 28 (18.4%) | - | - |  |
| n (% of column total) |  |  |  |  |
| **AIS Discharge**<br>Out of total n=137<sup>†</sup> |  |  |  |  |
| **AIS A** | 56 (40.9%) | 56 (80.0%) | - | **0.0001\*\*\*\*** |
| **AIS B** | 15 (10.9%) | 7 (10.0%) | 8 (20.0%) |  |
| **AIS C** | 12 (8.8%) | 7 (10.0%) | 5 (12.5%) |  |
| **AIS D** | 54 (39.4%) | - | 27 (67.5%) |  |
| n (% of column total) |  |  |  |  |
| **Neurological level**<br>Out of total n=152<sup>†</sup> |  |  |  |  |
| **C1-C4** | 68 (44.7%) | 23 (34.3%) | 22 (55.0%) | **0.0345\*** |
| **C5-C8** | 30 (19.7%) | 14 (20.0%) | 9 (22.5%) |  |
| **T1-T6** | 25 (16.4%) | 18 (25.7%) | 5 (12.5%) |  |
| **T7-T12** | 25 (16.4%) | 14 (20.0%) | 2 (5.0%) |  |
| **L1-S4** | 4 (2.6%) | 1 (1.4%) | 2 (5.0%) |  |
| n (% of column total) |  |  |  |  |
| **Multi-trauma**<br>n (% of column total) | 75 (46.0%) | 42 (60.0%) | 14 (35.0%) | **0.0170\*** |
| **NISS**<br>Median score (IQR) | 29.0 (21.0-38) | 38.0 (29.0-43.0) | 24.0 (20.0-32.3) | **<0.0001\*\*\*\*** |
| **Cause of injury** |  |  |  |  |
| **Transport accidents** | 85 (52.1%) | 41 (58.6%) | 18 (45.0%) | **0.0104\*** |
| **Falls** | 49 (30.1%) | 18 (25.7%) | 12 (30.0%) |  |
| **Diving / water sport** | 20 (12.3%) | 3 (4.3%) | 9 (22.5%) |  |
| **Other** | 9 (5.5%) | 8 (11.4%) | 1 (2.5%) |  |
| n (% of column total) |  |  |  |  |
| **Surgery**<br>n (% of column total) | 129 (79.1%) | 65 (92.9%) | 28 (70.0%) | **0.0023\*\*** |
| **Intubated on arrival**<br>n (% of column total) | 39 (23.9%) | 19 (27.1%) | 11 (27.5%) | >0.9999 |
| **Respiratory Infection**<br>n (% of column total) | 45 (27.6%) | 21 (30.0%) | 13 (32.5%) | 0.8319 |
| **Days in Hospital**<br>Median days (IQR) | 102.5 (39.5-211.3) | 192.5 (101.3-238.8) | 119 (76.50-216.8) | 0.0696 |

<sup>†</sup>Indicative of total sample size when excluding patients with missing AIS grading data. Patients with missing AIS data at admission and/or discharge, and patients graded with an admission AIS grade of D were excluded from AIS grade conversion analyses.
Patient groups with and without an AIS grade conversion were compared using the following statistical tests: Mann-Whitney U test for age, NISS and length of stay; Fisher's exact test for frequency statistics of variables with two possible outcomes, or $\chi^2$ test for variables with more than two possible outcomes. Categorical data are reported as absolute frequency and percentage, and metric variables (age, NISS and length of stay) as median with interquartile range (IQR). Abbreviations: AIS = American Spinal Cord Injury Association impairment scale; IQR = interquartile range; NISS = new injury severity score.

**Table 2:**
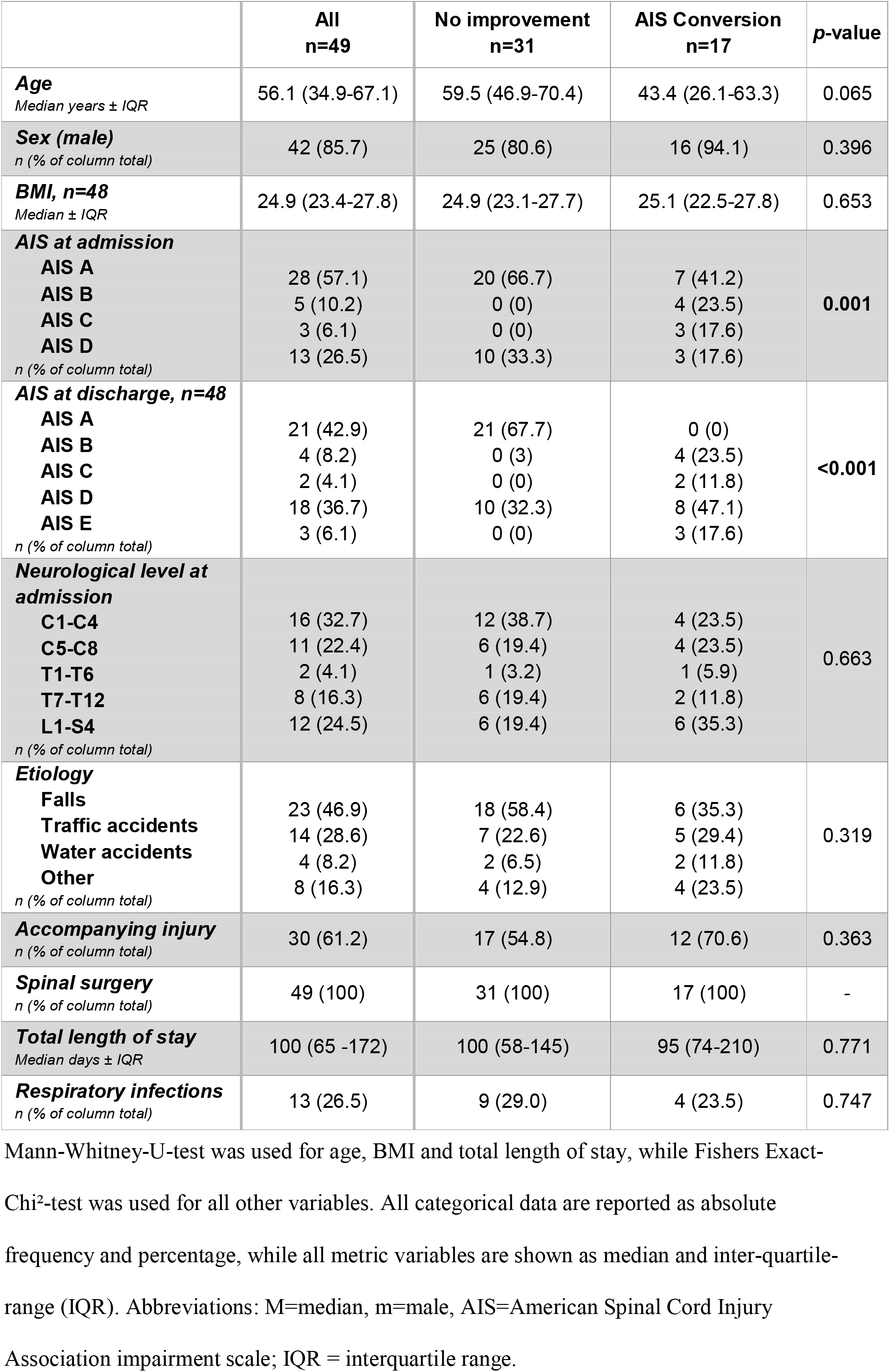
Baseline characteristics of the Berlin patient cohort

### Circulating WBC counts

For the Brisbane cohort, blood data was obtained from routine clinical care samples taken on admission (day 0), 1, 3 and 7 days post-injury (dpi). Peripheral venous blood samples were similarly collected for the Berlin cohort on admission, 3 and 8 dpi (*Supplementary* e-*Table 1A*). Absolute numbers of neutrophils, lymphocytes and monocytes within patient samples were measured at certified diagnostic laboratories of Pathology Queensland (Australia) and Labor Berlin GmbH (Germany), respectively. These counts were also used to calculate differences between timepoints, neutrophil-lymphocyte, neutrophil-monocyte, and monocyte-lymphocyte ratios for individual patients. Standard clinical reference ranges for differential WBC counts were as follows: 2.0-8.0×10^9^ neutrophils/L, 0.1-1.0×10^9^ monocytes/L, and 1.0-4.0×10^9^ lymphocytes/L in the Brisbane cohort, and 1.5-7.7×10^9^ neutrophils/L, 0.1-0.9×10^9^ monocytes/L, and 1.1-4.5×10^9^ lymphocytes/L in the Berlin cohort; the upper and lower limits for each reference range represent values that are two standard deviations away from the average of healthy controls (95% confidence interval). To increase the sensitivity of analysis, various WBC profiles were defined by categorical variables (i.e. ‘neutrophilia’ or ‘lymphopenia’) for select time points as specified. Clinical neutrophilia was defined as circulating neutrophil numbers exceeding the upper limit of the normal reference range (Brisbane-cohort >8.0 x 10^9^ cells /L; Berlin-cohort >7.7 x 10^9^ cells /L). Clinical lymphopenia was defined by a decrease in circulating lymphocyte numbers to below the normal reference range (Brisbane-cohort <1.0 x 10^9^ cells/L; Berlin-cohort <1.1 x 10^9^ cells /L).

### Injury severity and neurological assessment

For each patient, the extent of neurological impairment was assessed using the International Standards for Neurological Classification of Spinal Cord injury (ISNCSCI) / American Spinal Injury Association (ASIA) Impairment Scale (AIS). All patients were graded during the first week of admission to hospital (baseline). AIS conversion was used as a measure to assess recovery, where patients either converted to an improved AIS grade at discharge compared to their admission scores, or showed no improvement in that they received the same (or a worse) AIS grade upon leaving the hospital. Patients who were graded AIS D on admission were not included in any analyses that involved an assessment of patient AIS conversion rates due to ceiling effects with a limited subset sample size,^22^ except for an initial sensitivity analysis for all patients in the Berlin cohort (*Supplementary e-Table 2, 3 and 5*). The New Injury Severity Scale was used to provide a measure of the general extent of trauma sustained.^23^

### Respiratory infection and pneumonia classification

To validate the incidence of respiratory tract infections, including pneumonia, a combination of clinical tests and patient records were assessed together. Specifically, chest radiograph records were reviewed for indications of consolidation or opacity in the lungs, and this was then combined with sputum microbiology tests and clinician records of patient symptoms consistent with infection presentation. Designation of patients as being either with or without respiratory tract infections was performed prior to any further analysis. For the Berlin cohort, respiratory infections were further classified as being either tracheobronchial infections, defined in accordance with the Centres for Disease Control and Prevention (CDC) criteria,^24^ or ‘pneumonia’ with radiological findings based on CDC-criteria modified in accordance with recent consensus definitions for a ‘probable pneumonia’ or ‘definite pneumonia’ after stroke.^25^

### Statistical Analysis

For all graphs, continuous variables are presented as the median ± interquartile range (IQR), plotted in box and whisker plots with whiskers and outliers determined by the Tukey method. Categorical variables are presented as percentages. All statistical tests were two-tailed, with a p-value of less than 0.05 being considered statistically significant.

For the derivation (i.e. Brisbane) cohort, Fisher’s exact test was used to explore possible associations between categorical variables (i.e. neutrophilia, lymphopenia, lesion level, AIS conversion, presentation of respiratory infection, or intubation). For categorical variables with more than two outcomes in Table 1, a χ^2^ test was used. Mann-Whitney U test (exact) was used for analysis of data between two continuous variables, and a one-way ANOVA with Bonferroni’s *post hoc* test for comparison between multiple groups. Pearson’s correlation test was used to explore the statistical relationship between two continuous variables. Logistical regression modelling was employed to assess the incidence of AIS conversion in SCI patients, using neutrophilia (on day 0 and/or day 1) and lymphopenia (on day 1 and/or 3) as the predictor variable in separate models. These models were also subsequently adjusted for patient age, sex, the neurological level of the lesion, associated multi-trauma (MT), infection presentation as well as either NISS or admission AIS grade; using only one of the latter two covariates avoids potential over-adjustment of the model as the NISS already includes a rating of SCI severity^26^, meaning that it is not a fully independent parameter of AIS grade. The predictive power of the adjusted neutrophilia and lymphopenia models was further demonstrated with a Receiver Operating Characteristic (ROC) curve and Area Under the Curve (AUC) analysis. General linear modelling was used to test for an association between the incidence of respiratory infections and neutrophil-lymphocyte ratios at days 3 and 7 post-SCI. All statistical modelling was performed using SAS software (version 9.4, SAS Institute Inc.). All other statistical tests and visualisation of data was performed using GraphPad Prism (version 8.2; GraphPad Software, La Jolla, CA).

For the validation (i.e. Berlin) cohort, possible patient selection bias was examined first using a logistic regression model with the availability of WBC counts as the dependent variable and other inclusion criteria and relevant patient characteristics (AIS grade at admission, neurological level of injury, age, BMI, sex and accompanying injury) as independent variables (*Supplementary e-Table 1B*).^27^ No evidence for selection bias was found based on the relatively small sample of SCI patients with available WBC data, nor was there any association for any of the baseline parameters with an increased risk of exclusion from analysis. Linear mixed effects models with random intercept were then used to describe temporal changes for log transformed WBC data relative to the time between injury and blood collection; all independent variables were used as fixed effects. For statistical comparisons regarding neurological recovery, the study population was divided into two groups by AIS conversion (i.e. no improvement vs. improvement in AIS grading). For explorative analysis, patient groups were compared using the Mann-Whitney U test for continuous variables, or a χ^2^ test for categorical variables reporting the exact significances. To adjust for the risk of a non-improvement in AIS grade being due to possible confounders, logistic regression models were used with AIS grade conversion as the dependent variable and neutrophil numbers the covariate. Regression results were calculated both unadjusted and once adjusted for AIS grade at admission, the neurological level of injury at admission, age, BMI, sex, and presence of an accompanying injury. As a sensitivity analysis for neutrophilia effects, we also conducted the logistic regression models with neutrophilia categories as a covariate instead of continuous numbers. Neutrophilia was defined by neutrophil counts being above the reference value within the first 3 days after SCI.

Associations of respiratory infections with clinical and demographic factors such as AIS grading, neurological level of injury, mechanical ventilation, neutrophilia or neutrophil-lymphocyte ratios were analyzed in descriptive manner. Temporal differences in neutrophil-lymphocyte ratios between patients with and without respiratory infections were additionally analyzed using linear mixed effects models with random intercept, with all independent variables treated again as fixed effects. Statistical analyses were performed in the COaT-SCI database version as of September 6, 2019 using the software SPSS (Version 25.0, IBM, Armonk, NY, USA).

### Data Availability Statement

Requests for access to study data should be directed to the corresponding authors for consideration, and can be provided pending appropriate institutional review board approvals.

## RESULTS

### SCI patient cohort details and characteristics

The selection and analysis chart for the Brisbane patient cohort is shown in *Figure 1*. Out of the 163 patients with a traumatic SCI that were screened, a total of 2 individuals were excluded because of a lack of WBC data. Selection for inclusion of the remaining 161 patients was based on the type of analysis being undertaken, and the availability of specific patient data required for this. The age of the Brisbane cohort was 40 ± 28 years (median ± IQR), and 85.3% were males. The heterogeneous nature of SCIs was evident from individual differences in AIS grading, the neurological level of injury, cause of injury, and also the overall trauma severity for each patient as reflected in their NISS. The patient data presented in *Table 1* was segregated based on those individuals who either showed a documented AIS grade conversion indicative of neurological recovery (36.4%), or those with no improvement between admission and discharge (63.6%). Age, sex, and frequency of intubation on arrival were not different between these two subgroups of patients (p>0.59); a statistically non-significant trend towards a greater length of stay in hospital was observed for SCI patients with no AIS grade conversion (p=0.0696). The proportion of patients who acquired a respiratory infection was also not different, with 30.0% of patients with no substantial neurological recovery acquiring such an adverse event compared to 32.5% of patients that did show an AIS grade conversion (p=0.8319). We did observe an influence of SCI severity, the neurological level of injury, multi-trauma, NISS, cause of injury, and the need for surgery (p<0.05 for all variables) on longer-term neurological outcomes, i.e. when comparing individuals with a positive AIS grade conversion between admission and discharge, and those with no improvement. The consort diagram for the Berlin cohort is shown in *Supplementary e-Figure 1*, and the baseline characteristics of the patients are shown in *Table 2*. The relationship between AIS grade and conversion was also found for this patient cohort (p<0.001); patients with AIS conversion tended to be slightly younger in age compared to those patients who showed no neurological improvement (p=0.065).

**Figure 1:**
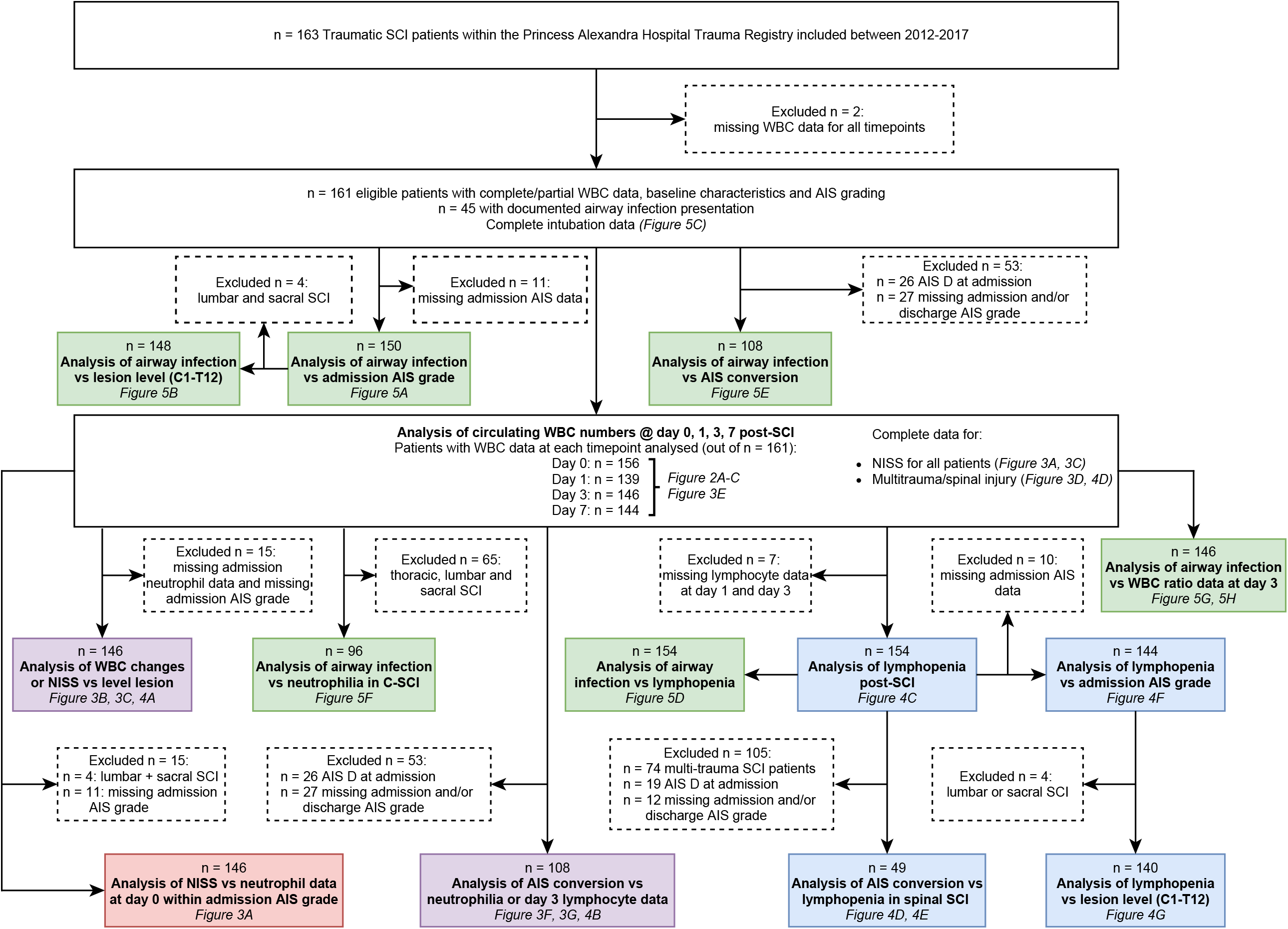
Diagram for the Brisbane exploration cohort. Available SCI patients were screened for eligibility based on the availability of WBC data. Patient datasets used for specific statistical analyses depended on the subgroup being tested, sample size limitations and/or availability of data; the resulting exclusion of patients for each subgroup analysis is indicated in the dashed boxes. The colour coding for each of the analysis boxes refers to the specific figure it pertains to, i.e. *red* = neutrophil-related (Figure 3); *blue* = lymphocyte-related (Figure 4); *purple* = either neutrophil or lymphocyte-related (Figure 3 or 4); *green* = respiratory infection-related (Figure 5). Abbreviations: AIS = American Spinal Cord Injury Association Impairment Scale; NISS = new injury severity score; SCI = spinal cord injury; WBC = white blood cell; C1-T12 = Cervical level 1 - Thoracic level 12; C-SCI = cervical spinal cord injury.

### Changes in circulating white blood cells during the acute phase of spinal cord injury

To gain a better appreciation of the systemic response to a traumatic SCI, we next plotted the circulating neutrophil, monocyte and lymphocyte counts in patient blood samples from admission (day 0), as well as 1, 3 and 7 days post-SCI for the Brisbane cohort (*Figure 2A-C*), and 0, 3 and 8 days post-SCI for the Berlin cohort (*Figure 2D-F*).

**Figure 2:**
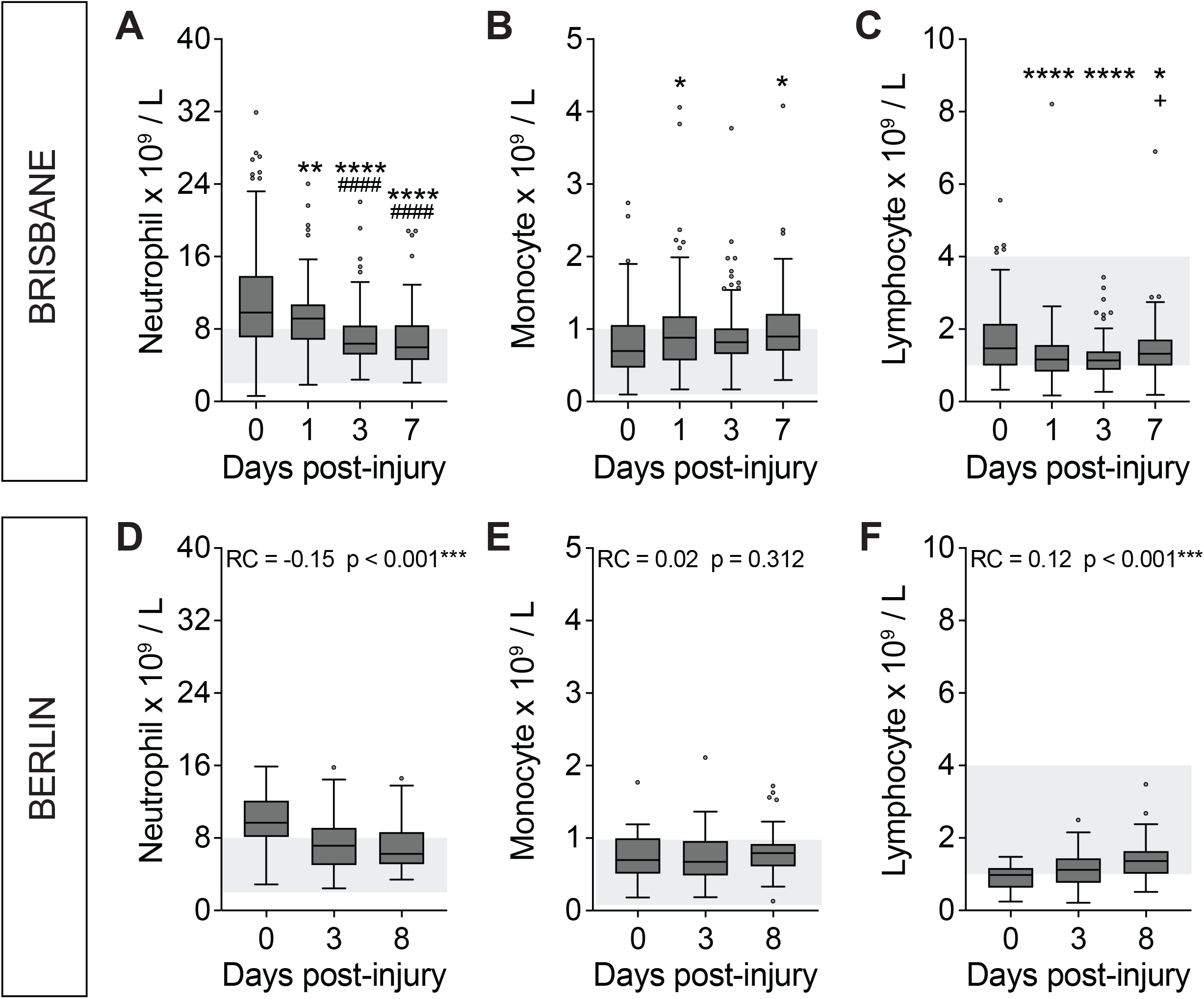
The systemic response to traumatic SCI is hallmarked by acute neutrophila and lymphopenia. For the exploration (i.e. Brisbane) cohort **(A-C)**, changes in circulating WBC numbers were compared between the specified timepoints (admission (0), 1, 3 and 7 days post-injury (dpi)) using one-way ANOVA with Bonferroni’s *post hoc* test. For the validation (i.e. Berlin) cohort **(D-F)**, temporal changes in circulating WBC numbers between 0 and 8 dpi were compared using a linear mixed effects model, with the indicated significances referring to the outcome of these models. **(A, D)** Circulating neutrophil numbers were significantly elevated above the clinical reference range (indicated in light grey) at admission and/or 1 dpi for both the Brisbane and Berlin patient cohorts and then rapidly decreased over time. **(B, E)** Circulating monocyte numbers were significantly increased at 1 and 7 dpi in the Brisbane cohort (relative to admission), but not between 1, 3 and 7 dpi; no difference was seen here within the Berlin cohort either. **(C, F)** Circulating lymphocyte numbers in the overall Brisbane cohort were significantly reduced at 1 and 3 dpi compared to admission; this subclinical lymphopenia was mostly resolved by 7 dpi. A clear lymphopenia was also evident for the Berlin patient group at 1 and 3 dpi, along with an upturn in lymphocyte numbers by 8 dpi. Box and whisker plots are shown with the median and interquartile range, Tukey whiskers, and outliers indicated as dots. Normal clinical reference ranges for each of the main WBC types were: neutrophils = 2 – 8 x 10^9^ cells/L; monocytes = 0.1-1 x 10^9^ cells/L; lymphocytes = 1 – 4 x 10^9^ cells/L in the Brisbane cohort and: 1.5-7.7 x 10^9^ neutrophils/L,0.1-0.9 x 10^9^ monocytes/L, and 1.1-4.5 x 10^9^ lymphocytes/L in the Berlin cohort. *p<0.05, **p<0.01, ****p<0.0001 (compared to admission / 0 dpi); ^####^p<0.0001 (compared to 1 dpi); ^+^p<0.05 (compared to 3 dpi). RC = regression coefficient (95% confidence interval).

Circulating neutrophil numbers for patients in the Brisbane cohort were acutely elevated in response to SCI and typically well above the normal range at the time of admission to hospital (day 0) and at 1 dpi (*Figure 2A*). Although the neutrophil count typically started to fall between 0 and 1 dpi (p=0.0081), their numbers usually remained above clinical reference range for the first 24 hours after injury. This immediate SCI-induced neutrophilia within the peripheral blood typically resolved itself during the first week, as evident from the fact that the circulating neutrophil count was significantly lower again and mostly within the normal range by 3 and/or 7dpi (p<0.0001 compared to both admission and 1 dpi; *Figure 2A*). Monocyte counts mostly remained within the clinical reference range during the first week post-SCI, although some temporal changes were observed in that their numbers were significantly increased at 1 dpi (p=0.0351), then dropped on 3 dpi (p=0.5719), before rising again at 7 dpi (p=0.0108 relative to admission/day 0; *Figure 2B*). Lastly, circulating lymphocyte numbers also mostly remained within the normal reference range for this patient cohort as a whole although a significant reduction was observed at 1 and 3 dpi compared to admission bloods (p<0.0001), signifying a subclinical lymphopenia that resolved itself by 7 dpi (*Figure 2C*).

Data from the Berlin cohort independently verified these observations, with circulating neutrophil numbers being significantly elevated above the normal reference range on day 0 post-SCI and then significantly decreasing over time until day 8 (p<0.001, regression coefficient (RC) (95% CI) = −0.15 (−0.22 – −0.07); *Figure 2D*). Monocyte numbers fluctuated modestly over time and, similarly to the Brisbane cohort, were highest on day 8 (i.e. ∼1 week post-SCI; *Figure 2E*), however no overall significant effect was observed (p=0.312, RC (95% CI) = 0.02 (−0.02 – 0.06)). Lymphocyte counts for the Berlin cohort were clearly below the normal reference range acutely after SCI (day 0) and then, similar to the Brisbane cohort, they mostly recovered during the first week post-injury (p<0.001, RC (95% CI) = 0.12 (0.07 – 0.16); *Figure 2F*). In summary, these data show that there is an elevated neutrophil response (neutrophilia) immediately after SCI, alongside a more subtle biphasic change in the circulating monocyte count, with an overall increase over time (i.e. between admission and 7 dpi), and a reduction of lymphocytes (subclinical and/or clinical lymphopenia) during the first week of hospitalization after SCI for both the Brisbane and Berlin patient cohorts.

### Circulating neutrophil numbers correlate with trauma severity

We next explored relevant factors that were likely to influence the magnitude of the acute neutrophil response to SCI. We first considered NISS, which rates the three most severe anatomic injuries for a given traumatic accident. NISS data was found to be positively correlated with circulating neutrophil numbers on admission (Pearson’s r^2^=0.09165, p<0.001, *Figure 3A*). Sub-analysis of NISS data showed that this relationship was mostly driven by SCI patients that were ‘complete’ (i.e. individuals with a complete loss of motor and sensory function below the level of injury (AIS grade A; Pearson’s r^2^=0.2234, p<0.0001) compared to those that were ‘incomplete’ (AIS grade B, C or D; Pearson’s r^2^=0.001), suggesting some influence of SCI severity / the neurological impairment at the time of the accident over the magnitude of the neutrophil response at the level of the blood.

**Figure 3:**
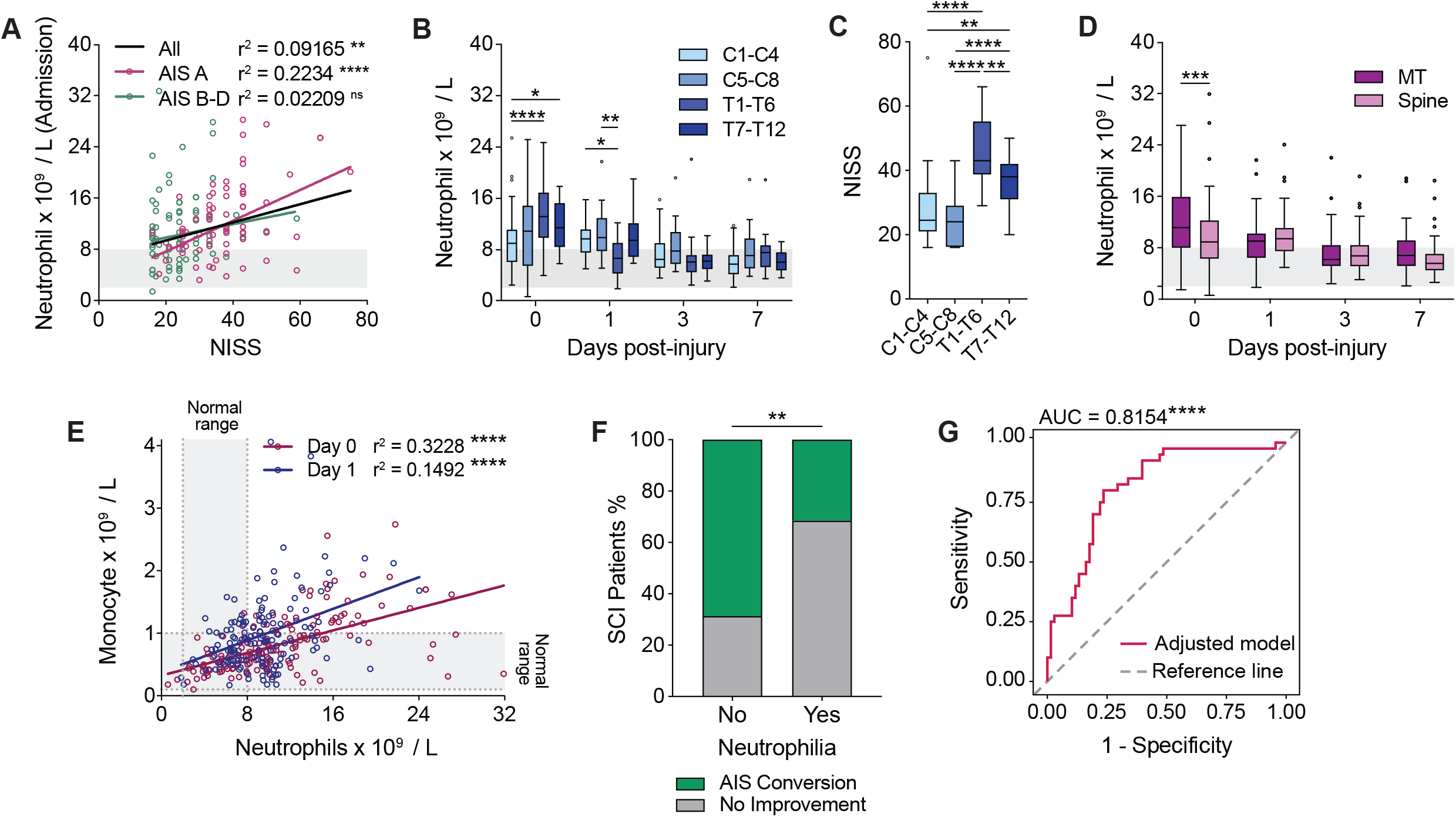
Circulating neutrophil numbers are associated with injury severity and long-term patient outcomes. **(A)** Circulating neutrophil numbers on admission positively correlate with the trauma severity as indicated by NISS data. This association was also present in patients with an admission AIS grade of A (complete SCI) versus those with an AIS grade of B, C or D (incomplete SCI); r^2^ value = Pearson’s correlation coefficient; linear fit indicated by solid black line for all patients, dashed lines for subgrouping based on admission AIS grade. **(B)** Blood neutrophil numbers during the first week of SCI segregated by lesion level (two-way ANOVA with Bonferroni’s *post hoc* test) **(C)** NISS data for patients with cervical versus thoracic injuries (one-way ANOVA with Bonferroni’s *post hoc* test). **(D)** SCI patients with associated multi-trauma (MT) had significantly higher circulating neutrophil numbers at admission compared to those with an spine-isolated spinal cord injury (SCI; two-way ANOVA with Bonferroni’s *post hoc* test). **(E)** Circulating neutrophil numbers are positively correlated with circulating monocyte numbers at admission (purple) and 1 day post-injury (blue). The reference range for each WBC type is indicated in light grey and on the relevant axis of the graph (r^2^ value = Pearson’s correlation coefficient; linear fit is indicated by the solid lines). **(F)** AIS grade conversion rates in patients with and without acute neutrophilia (Fisher’s exact test). **(G)** ROC curve analysis of the adjusted neutrophilia model used to predict AIS conversion showed high specificity and sensitivity as indicated by a significant AUC (red line = adjusted model, dashed grey line = reference line where AUC = 0.5). Bar graphs in B-D are box and whisker plots showing the median and interquartile range, and Tukey whiskers; outliers are indicated by dots. Clinical reference ranges for WBC types are shown in light grey. AIS = American Spinal Cord Injury Association Impairment Scale; AUC = area under the curve; C = cervical; NISS = new injury severity score; ROC = receiver operating characteristic; SCI = spinal cord injury; T = thoracic; WBC = white blood cell. *p<0.05, **p<0.01, ***p<0.001, and ****p<0.0001.

When segregating patients based on the neurological level of their injury, we found that there was usually a more pronounced acute neutrophilia (day 0) in individuals with a thoracic SCI compared to those with cervical injuries (*Figure 3B*). This finding was particularly prominent in patients with an upper thoracic SCI (T1-T6 vs. C1-C4: p<0.0001; T7-T12 vs. C1-C4: p<0.05) and is most likely related to the significantly higher NISS for this subgroup compared to those with cervical SCIs (p<0.0001; *Figure 3C*). Consistent with this, and the view that traumatic injuries involving the cervical spine require less force to cause an SCI compared to the thoracic region (where the vertebral canal space is typically also narrower ^28^), patients with thoracic SCIs were also much less likely to have a positive conversion in their AIS grade during their recovery (p=0.0026; *Supplementary e-Figure 2A*). The influence of trauma severity on the neutrophil response was also evident in multi-trauma (MT) patients who had significantly higher neutrophil numbers upon admission compared to those with an isolated spinal injury (p=0.0001; *Figure 3D*). Taken together these results corroborate the recognised link between neutrophil mobilisation and traumatic injuries, including in SCI patients.

### A greater neutrophil response immediately after SCI is associated with poorer prospects for long-term functional recovery

We next investigated how the systemic inflammatory response to SCI, in particular the extent of neutrophilia, correlated with neurological outcomes. For this, we used the available data from AIS grading, which is the universally accepted assessment tool to assess neurological impairment in SCI patients and performed routinely upon admission / in the first week of SCI (baseline) and then again at discharge.^1^ We recently already reported that patients who convert to an improved AIS grade at discharge typically have significantly lower neutrophil numbers on admission.^8^ Further examination of this Brisbane cohort of patients revealed a diverging neutrophil profile, where the majority of individuals presented with clinical neutrophilia on admission and/or day 1 (79.5% of the study cohort), except for a smaller subgroup of patients (20.5%) in which neutrophil numbers at all times remained within the normal reference range during the entire first week post-accident (hereafter referred to as “within range”; *Supplementary e-Figure 2B*). Patients that displayed acute clinical neutrophilia usually also had elevated monocyte counts compared to those that were classed as “within range” (0 dpi: p=0.0046; 1dpi: p=0.0672; *Supplementary e-Figure 2C*). Accordingly, circulating neutrophil numbers were positively correlated with monocyte counts in blood samples from both admission and day 1 post-SCI (*Figure 3E*), suggesting a joint mechanism that drives the activity of these WBCs in a patient-specific manner. We then further probed the putative significance of the systemic neutrophil response relative to SCI outcomes based on whether patients presented with acute neutrophilia or stayed within range. In doing so, we uncovered that a significantly greater proportion of SCI patients where neutrophil numbers stayed within the normal reference range converted to an improved AIS grade (*Figure 3F*). This association remained in an adjusted logistical regression model where age, sex, lesion level, associated multi-trauma and NISS were accounted for (Effect of neutrophilia on non-conversion: OR (95% CI) = 4.16 (1.13 – 15.30), p=0.0317; *Table 3A*), or when admission AIS grade was used instead of NISS (Effect of neutrophilia on non-conversion: OR (95% CI) = 5.886 (1.52 – 22.83), p=0.0104). ROC curve was used to assess the predictive power of this model, which revealed a significant Area Under Curve (AUC) indicative of high sensitivity and specificity (AUC (95% CI) = 0.8154 ± 0.0418 (0.7334 – 0.8975), p <0.0001; *Figure 3G*). We lastly also explored here whether the temporal trajectory of neutrophilia resolution was a factor in the likelihood of AIS grade conversion. Using an adjusted logistical regression model with patient age, sex, lesion level, lesion severity (admission AIS grade), multi-trauma and infection presentation during the first week post-injury all accounted for, we found no evidence for a correlation or influence of the decrease in neutrophil numbers between days 0 and 1 and patient outcomes (Effect of neutrophil decrease on AIS grade conversion, day 0-1: OR (95% CI) = 1.08 (0.90 – 1.29), p=0.417; *Supplementary e-Table 4A and Supplementary e-Figure 2D*). We did observe a relationship for the calculated difference in neutrophil numbers between days 1 and 3, with a greater decrease being more likely associated with AIS grade conversion failure rather than improvement (Effect of neutrophil decrease on AIS grade conversion, day 1-3: OR (95% CI) = 0.765 (0.635 – 0.921), p=0.0047; *Supplementary e-Table 4B and Supplementary e-Figure 2E*). This finding is consistent with the extent of acute neutrophilia affecting outcomes, and also the calculated decrease during the early stages of neutrophilia resolution being very much influenced by the admission counts. On the other hand, a greater decrease during the sub-acute phase (days 3-7 post-injury) was associated with an increased likelihood of AIS grade conversion (Effect of neutrophil decrease on AIS grade conversion, day 3-7: OR (95% CI) = 1.556 (1.166 – 2.077), p=0.0027; *Supplementary e-Table 4C and Supplementary e-Figure 2F*).

**Table 3:** Association of neutrophilia and the neurological outcome from SCI

**3A: Brisbane cohort**
|  | Unadjusted OR | 95% CI | p-value |
| --- | --- | --- | --- |
| *Neutrophilia, yes* | 4.778 | 1.521-15.012 | **0.0074** |
| Nagelkerke's R<sup>2</sup> | 0.0955 |  | n = 108 |
|  | Adjusted OR | 95% CI | p-value |
| *Neutrophilia, yes* | 4.164 | 1.133-15.303 | **0.0317** |
| *Age* | 1.019 | 0.990-1.048 | 0.1992 |
| *Sex, female* | 0.889 | 0.229-3.447 | 0.8645 |
| *NISS* | 0.904 | 0.842-0.971 | **0.0054** |
| *Level* |  |  | 0.7164 |
| *Cervical* | 1.407 | 0.394-5.022 | 0.7633 |
| *Lumbosacral* | 3.202 | 0.167-61.479 | 0.4854 |
| *Thoracic (reference)* | - | - | - |
| *Multi-trauma, yes* | 1.042 | 0.340-3.190 | 0.9432 |
| Nagelkerke's R<sup>2</sup> | 0.3610 |  | n = 108 |

**3B: Berlin cohort**
|  | Unadjusted OR | 95% CI | p-value |
| --- | --- | --- | --- |
| *Neutrophilia, yes* | 18.33 | 1.87-180 | **0.013** |
| Nagelkerke's R<sup>2</sup> | 0.385 |  | n = 28 |

Clinical neutrophilia was defined by an increase in circulating neutrophil numbers above the clinical reference range on admission and/or 1 dpi. The association between neutrophilia and AIS conversion was assessed in both the Brisbane (**A**) and Berlin (**B**) patient cohorts with an admission AIS grade of A, B or C for validation; patients with an AIS grade of D were excluded from this analysis to minimise ceiling effects. For the Brisbane cohort, logistic regression analysis was applied using AIS grade conversion as the dependent variable (conversion = 0, non-conversion = 1) and clinical neutrophilia as the primary predictor variable. The model was first calculated without adjustment, and then corrected for age, sex, NISS, the level of the lesion (i.e. cervical, thoracic or lumbosacral) and multi-trauma. For the Berlin cohort, a univariate logistic model was used with AIS conversion as the dependent variable (conversion = 0, non-conversion = 1) and clinical neutrophilia as the primary predictor variable. Abbreviations: 95% CI = 95% confidence interval; AIS=American Spinal
Cord Injury Association Impairment Scale; dpi = days post-injury OR = odds ratio; NISS = new injury severity score.

Although slightly delayed compared to the Brisbane cohort, an inverse relationship between the acute circulating neutrophil count (3 dpi) and the likelihood of longer-term neurological recovery was confirmed in the Berlin cohort *(Supplementary e-Table 2A,B)*. Despite the smaller sample size, the association between a high acute neutrophil count with a similar categorisation of neutrophilic patients and the risk of no improvement in AIS grading also remained in a univariate model (Effect of neutrophilia on non-conversion: OR (95% CI) =18.33 (1.87 – 180), p=0.013; *Table 3B*). Even with the inclusion of AIS grade D patients, which reduces the likelihood of a result due to ceiling effects, the association between neutrophilia and AIS grade conversion failure still remained (*Supplementary e-Table 3*). Together, these clinical findings from a derivation and independent validation cohort provide strong evidence for a negative association between the magnitude of the neutrophil mobilization response and the outcome from SCI, where patients with neutrophilia early after injury have a worse functional prognosis while those with a profile that stays within the normal clinical range from the outset typically show better long-term functional recovery.

### Lymphopenia is associated with better functional recovery from SCI

With a clear reduction in systemic lymphocyte numbers observed during the first week of SCI in both the Brisbane and Berlin cohort (Figure 2C and F, respectively), we next examined if and how the lymphocytic blood content acutely after SCI links in with trauma characteristics and functional outcomes. Given that circulating lymphocyte numbers troughed relatively early, i.e. at ≤3 dpi, we first tested whether there was an inverse association between trauma severity (NISS) and lymphocytes numbers for the Brisbane cohort at this timepoint, but no such correlation was found within this patient group as a whole (*Supplementary e-Figure 3A)*. Further exploration of the data did reveal a significant increase in blood lymphocyte numbers in patients with lower thoracic injuries (T7-T12 vs. C1-C4: p=0.0077; *Figure 4A*), but this was only observed in admission bloods and not maintained at later timepoints. This result may suggest that there is a neural basis to lymphocyte mobilisation acutely after injury that is influenced by the lesion level. Alternatively, a higher incidence of multi-trauma in this patient group could also be a factor, prompting us to group patients within the study cohort based on whether they presented with multiple traumatic injuries (i.e. MT), or an isolated spine injury only. The temporal pattern / change in blood lymphocyte numbers during the first week post-SCI was, however, not different between these two subgroups (p>0.55; data not shown). Interestingly, patients in the Brisbane cohort with an isolated spine injury and who converted to an improved AIS grade typically had lower lymphocyte numbers at 3 dpi (p=0.0105); no such relationship was observed in multi-trauma patients (p=0.99; *Figure 4B*).

**Figure 4:**
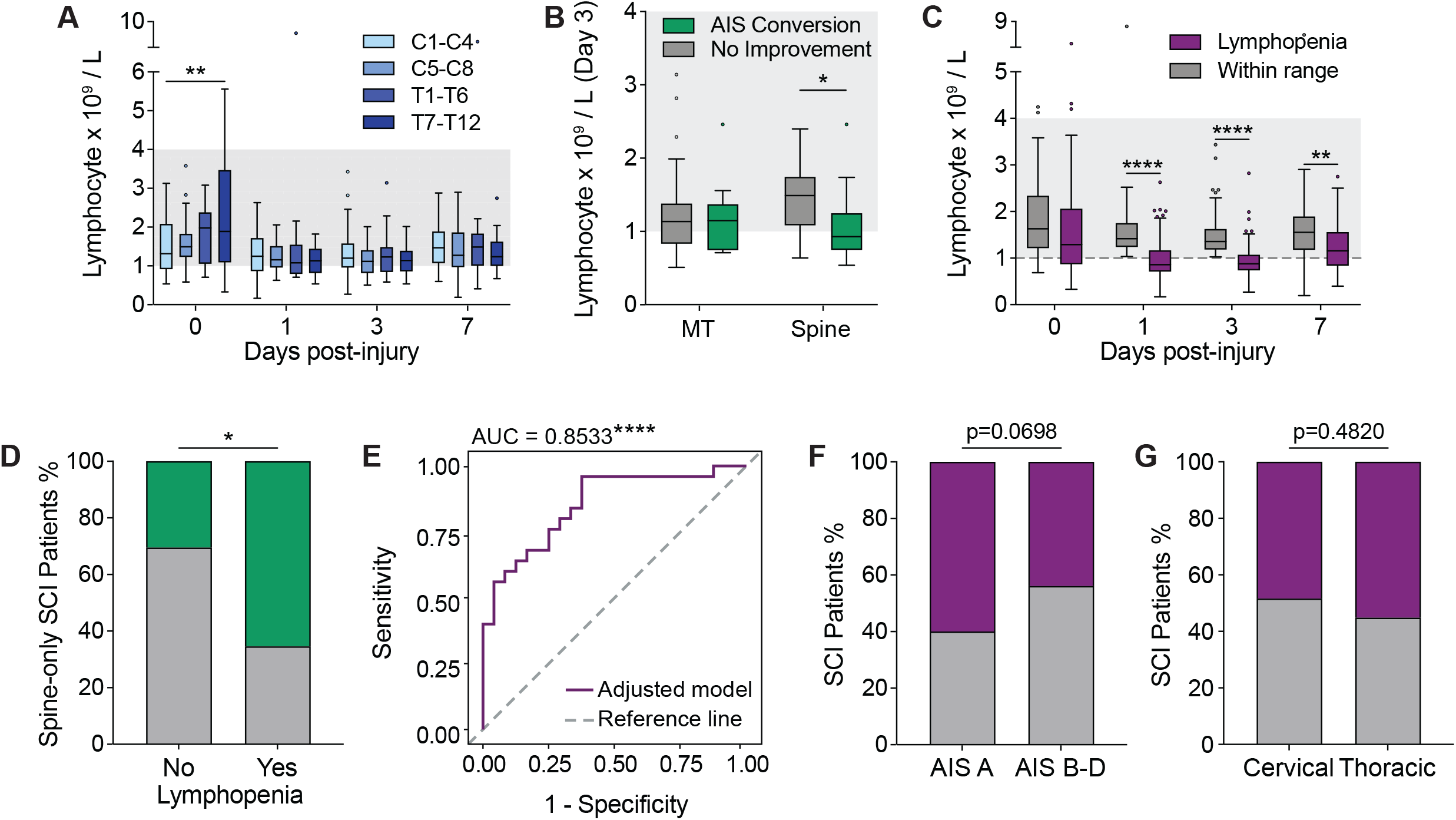
Lymphopenia is associated with better functional recovery in SCI. **(A)** Circulating lymphocyte counts segregated by lesion level during the first week post-SCI (two-way ANOVA with Bonferroni’s *post hoc* test); the clinical reference range is shown in light grey. **(B)** Lymphocyte numbers at 3 dpi in SCI patients with multi-trauma or an isolated spine injury, further segregated based on AIS grade conversion between admission and discharge (two-way ANOVA with Bonferroni’s *post hoc* test). **(C)** Sub-group analysis showing circulating lymphocyte numbers for patients with and without clinical lymphopenia; the dashed grey line indicates the lower reference range limit used to classify lymphopenia on day 1 and/or 3 post-SCI (two-way ANOVA with Bonferroni’s *post hoc* test). **(D)** AIS grade conversion rates in SCI patients with and without acute clinical lymphopenia portion (isolated spine injury only; Fisher’s exact test). **(E)** ROC curve analysis of the adjusted lymphopenia model used to predict AIS conversion showing specificity and sensitivity as indicated by a significant AUC (purple line = adjusted model, dashed grey line = reference line where AUC= 0.5). **(F)** Incidence of lymphopenia in patients with a complete SCI (AIS grade A) versus those with incomplete SCI (AIS grade B, C or D; Fisher’s exact test). **(G)** Incidence of lymphopenia relative to lesion level (Fisher’s exact test). Box and whisker plots in A-C show the median and interquartile range, Tukey whiskers, and outliers as dots. AIS = American Spinal Cord Injury Association Impairment Scale; AUC = area under the curve; dpi = days post injury; C = cervical; MT = multi-trauma; NISS = new injury severity score. ROC = receiver operating characteristic; SCI = spinal cord injury; T = thoracic. *p<0.05, **p<0.01, ****p<0.0001.

To further explore a putative link between lymphopenia and the longer-term neurological outcome, we segregated patients within the Brisbane cohort based on whether the lymphocyte numbers for individual patients dropped to below the clinical reference range at 1 and/or 3 dpi; these patients (52.6% of the cohort) will hereafter be referred to as the “lymphopenia” subgroup while the remaining patients (47.4% of the study cohort) were clustered as the “within range” subgroup (*Figure 4C*). Subsequent assessment of neurological recovery showed that patients with an isolated spine injury and clinical lymphopenia were significantly more likely to have improved functional outcomes and convert to a better AIS grade compared to those individuals where circulating lymphocytes numbers remained within range (p=0.022; *Figure 4D*). This result is supported by a logistical regression model correcting for age, sex, NISS and neurological lesion level (Effect of lymphopenia on conversion: OR (95% CI) = 24.15 (3.06 – 190.75), p=0.0025; *Table 4*), or those using admission AIS grade instead NISS (Effect of lymphopenia on conversion: OR (95% CI) = 35.99 (2.43 – 533.42), p=0.0092). Analysis of the ROC curve showed that the AUC was significant and the model thus has validity in terms of both specificity and sensitivity for predicting AIS conversion (AUC (95% CI)= 0.8533 ± 0.0542 (0.7472 – 0.9595), p <0.0001; *Figure 4E*). Lymphopenia itself was otherwise trending to more likely occur in patients that were classed as ‘complete’ (i.e. AIS grade A) on admission (p=0.0698, *Figure 4F*). We next tested whether the manifestation of lymphopenia in our study cohort was dependent on the level of injury and more common in those with high-level SCI, but did not observe such a link (p=0.4820; *Figure 4G*). If anything, a trend towards a greater prevalence of lymphopenia was found in patients with an injury below T6 (p=0.0601, *Supplementary e-Figure 3B*). Taken together, these results demonstrate that clinical lymphopenia is present in a subgroup of patients during the first week of SCI and that this phenomenon may be associated with more favourable long-term outcomes in the derivation (i.e. Brisbane) cohort. It should be noted, however, that we were not able to confirm this finding within the Berlin cohort, most likely because of a smaller sample size that did not allow for subgroup stratification (*Supplementary e-Table 5*). Further research is therefore required to independently validate this observation.

**Table 4:** Association of lymphopenia and neurological recovery in the Brisbane Cohort (within isolated spine injury subset)

|  | **Unadjusted OR** | **95% CI** | **p-value** |
| --- | --- | --- | --- |
| *Lymphopenia, yes* | 4.317 | 1.299-14.344 | **0.0170** |
| Nagelkerke's R<sup>2</sup> | 0.1561 |  | n = 49 |
|  | **Adjusted OR** | **95% CI** | **p-value** |
| *Lymphopenia, no* | 24.153 | 3.058-190.752 | **0.0025** |
| *Age* | 0.952 | 0.899-1.008 | 0.0917 |
| *Sex, female* | 0.747 | 0.082-6.786 | 0.7951 |
| *NISS* | 1.209 | 1.045-1.399 | **0.0109** |
| *Level* |  |  | 0.8256 |
| *Cervical* | 0.621 | 0.063-6.160 | 0.9353 |
| *Lumbosacral* | 0.326 | 0.009-11.774 | 0.5796 |
| *Thoracic (reference)* | - | - | - |
| Nagelkerke's R<sup>2</sup> | 0.4998 |  | n = 49 |

Clinical lymphopenia was defined by a decrease in circulating lymphocyte numbers below the clinical reference range at 1 and/or 3 dpi. The association between lymphopenia and AIS grade conversion was tested in all 'spine-only' SCI patients with an admission AIS grade of A, B or C; patients with an AIS grade of D were excluded from this analysis to minimise ceiling effects. Logistic regression analysis was applied using AIS conversion as the dependent variable (non-conversion = 0, conversion = 1) and clinical lymphopenia as the primary predictor variable. The model was calculated without adjustment first, and then again corrected for age, sex, NISS and level of the lesion (i.e. cervical, thoracic or lumbosacral). Abbreviations: 95% CI = 95% confidence interval; AIS=American Spinal Cord Injury Association Impairment Scale; dpi = days post-injury; OR = odds ratio; NISS = new injury severity score; SCI = spinal cord injury.

### Respiratory infections are correlated with neutrophilia in cervically injured patients

We lastly investigated the incidence of respiratory infections such as pneumonia for both the Brisbane and Berlin patient cohorts. For the Brisbane cohort, the presence of respiratory infections in SCI patients was based on records of symptom presentation, subsequent chest radiograph analysis and sputum culture results; the various clinical characteristics of these patients are described in *Table 5*. Fifty percent of these patients acquired their infection 1 ± 1 day after injury. Sputum microbiology tests revealed that *Staphylococcus, Candida* and *Pseudomonas* were among the most commonly cultured microbes (*Supplementary e-Figure 4A*), with *Staphylococcus aureus* being the predominant species. We found no association between the incidence of infection and whether patients had a complete or incomplete SCI, i.e. AIS grade A vs. B-D (p=0.5913; *Figure 5A*). These findings are comparable to observations in the Berlin cohort (p=0.6889; *Figure 5I*). Consistent with existing literature,^29^ the prevalence of respiratory infections was otherwise significantly higher in patients with a cervical SCI for both the Brisbane and Berlin cohorts (p=0.0066 and p=0.0246, respectively; *Figure 5B, J*). Patients that were intubated on arrival at the hospital or were requiring mechanical ventilation during acute care were also more likely to contract a respiratory infection (p=0.0355 and p=0.0078, respectively; *Figure 5C, K*), which agrees with clinical observations regarding ventilator-associated pneumonia.^29^

**Table 5:**
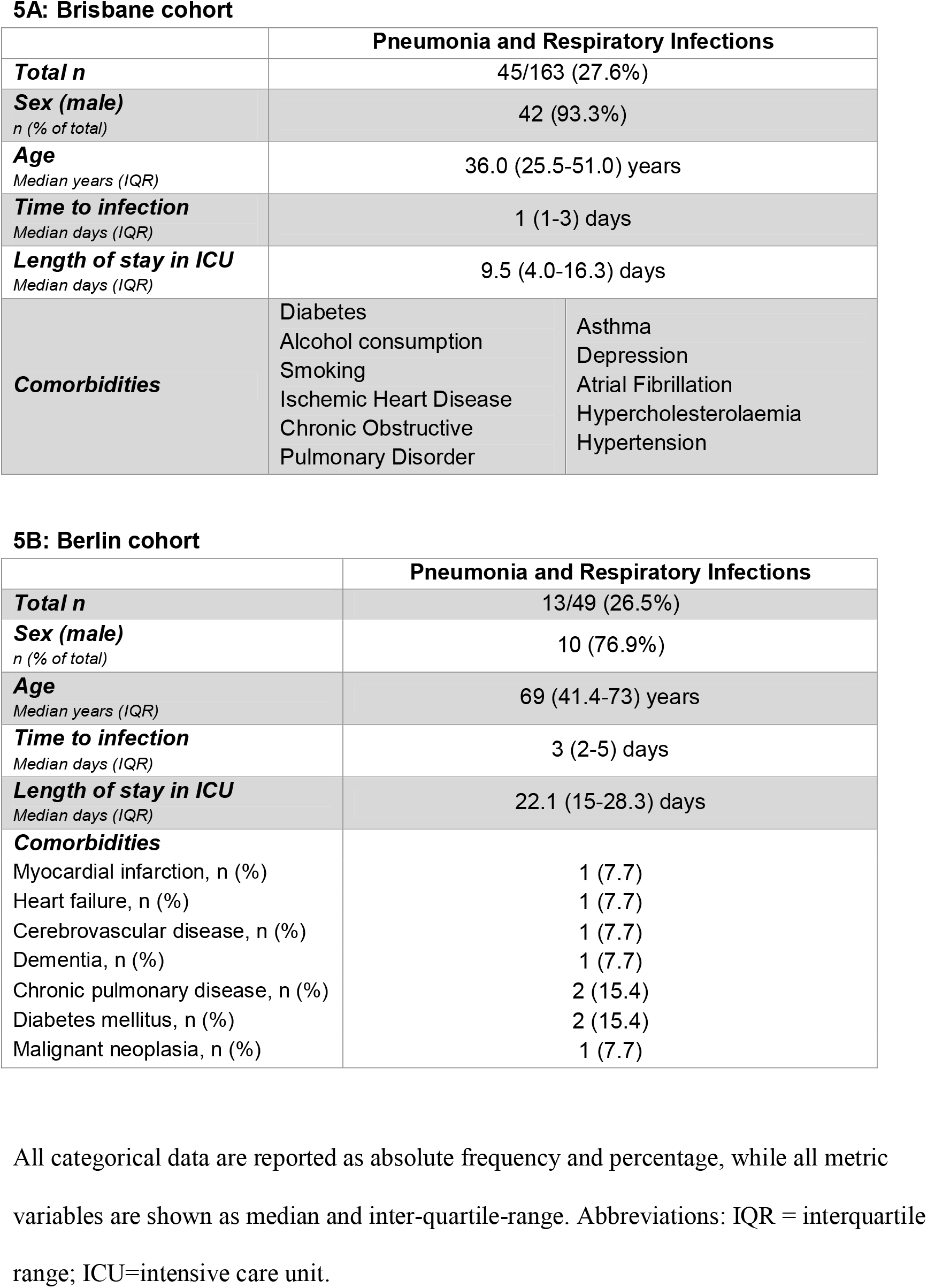
Clinical characteristics of patients with respiratory infections

**Figure 5:**
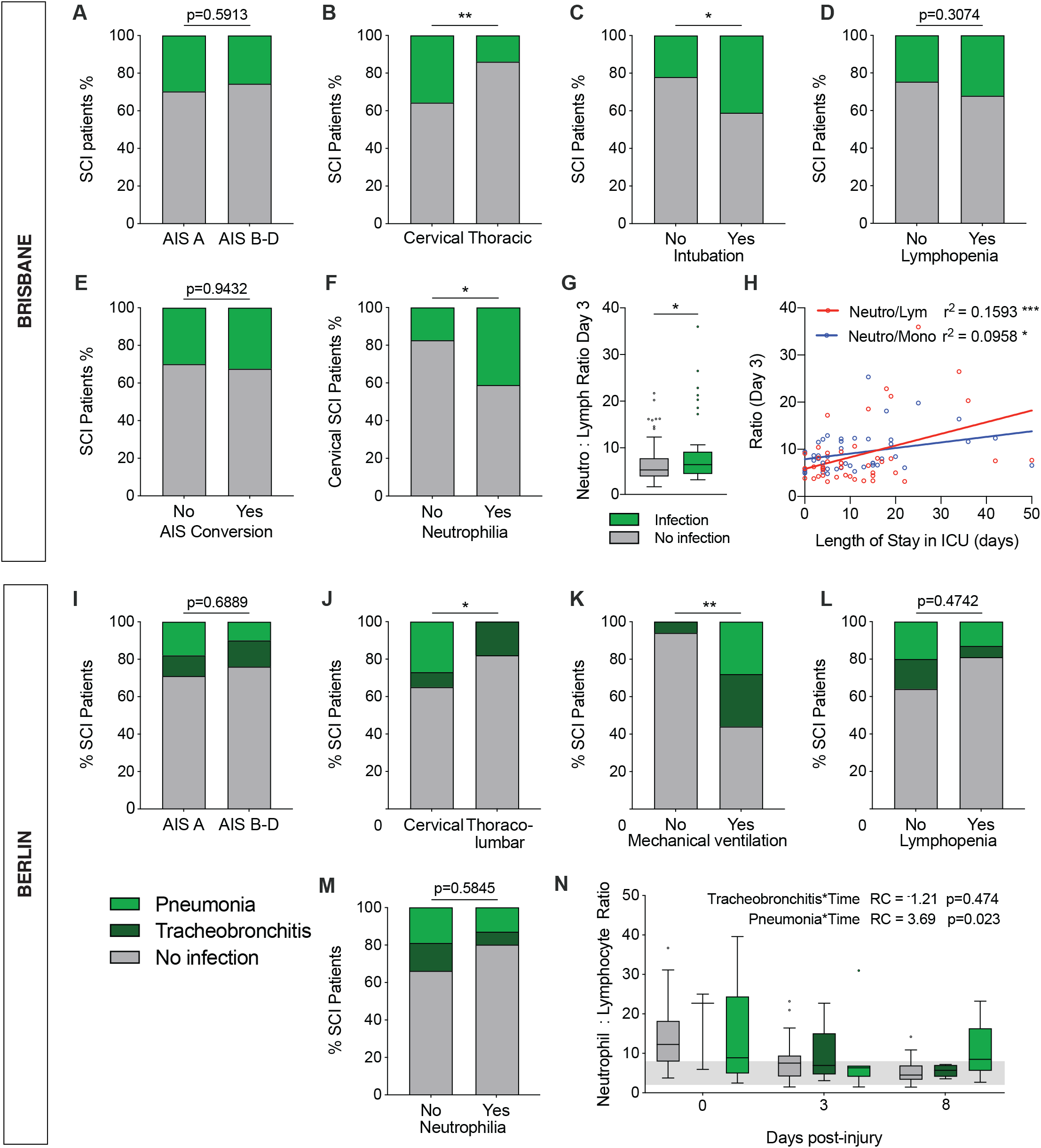
Respiratory infection presentation and clinical outcomes in SCI patients. Patients presenting with pneumonia or other respiratory infections acquired after injury were compared with their non-infected counterparts for white blood cell (WBC) changes, injury characteristics and neurological recovery in both the Brisbane **(A-H)** and Berlin **(I-N)** patient cohorts. **(A, I)** Incidence of respiratory infections in patients with complete SCI (AIS grade A) versus incomplete SCI (AIS grade B, C or D; Fisher’s and *χ*^2^ test for A and I, respectively). Patients with cervical injury **(B, J)** and those that were intubated / mechanically ventilated **(C, K)** were more likely to acquire an airway infection (Fisher’s and *χ*^2^ test for B-C and J-K, respectively). **(D, L)** Incidence of respiratory infections in SCI patients with and without acute lymphopenia (1 and/or 3 dpi; Fisher’s and *χ*^2^ test for D and L, respectively). **(E)** Incidence of respiratory infections in patients with and without an AIS grade conversion between admission and discharge. **(F, M)** Incidence of respiratory infections in cervically-injured patients with and without acute neutrophilia (Fisher’s and *χ*^2^ test for F and M, respectively). **(G, N)** Neutrophil-lymphocyte ratios for SCI patients with and without acquired airway infections (Mann-Whitney test for G; for linear mixed model estimates related to N see *Table 6B*). **(H)** Relationship between neutrophil-lymphocyte and neutrophil-monocyte ratios (3 dpi) to the length of stay in the intensive care unit (ICU; r^2^ value = Pearson’s correlation coefficient; linear fit indicated by solid coloured lines). Box and whisker plots show the median and interquartile range, Tukey whiskers, and outliers as dots. AIS = American Spinal Cord Injury Association Impairment Scale; NISS = new injury severity score; dpi = days post-injury; ICU = intensive care unit; WBC = white blood cell. *p<0.05, **p<0.01, ***p<0.001, ****p<0.0001.

To explore any potential link between infection susceptibility, lymphopenia and/or neutrophilia, we next compared WBC profiles of infected versus non-infected patients. No significant differences were observed in the circulating neutrophil, monocyte or lymphocyte count between these subgroups in the first week following SCI (Brisbane cohort; *Supplementary e-Figure 4B-D*). We also found no significant differences in infection presentation when comparing patients with and without lymphopenia (p=0.3074, *Figure 5D;* p=0.4742, *Figure 5L*), nor was there any significant association with AIS grade conversion (p=0.9432, *Figure 5E*), including when corrected for lesion level, or trauma severity (*Supplementary e-Figure 4E*). We did find that the incidence of respiratory infections was higher in the Brisbane cohort for patients that displayed a significant neutrophilia on admission, but this only applied to those with a cervical SCI (p=0.0467, *Figure 5F*) and not the study cohort as a whole (p=0.5116; *Supplementary e-Figure 4F*). A similar statistically non-significant association was observed in the Berlin cohort (*Figure 5M*).

**Table 6:**
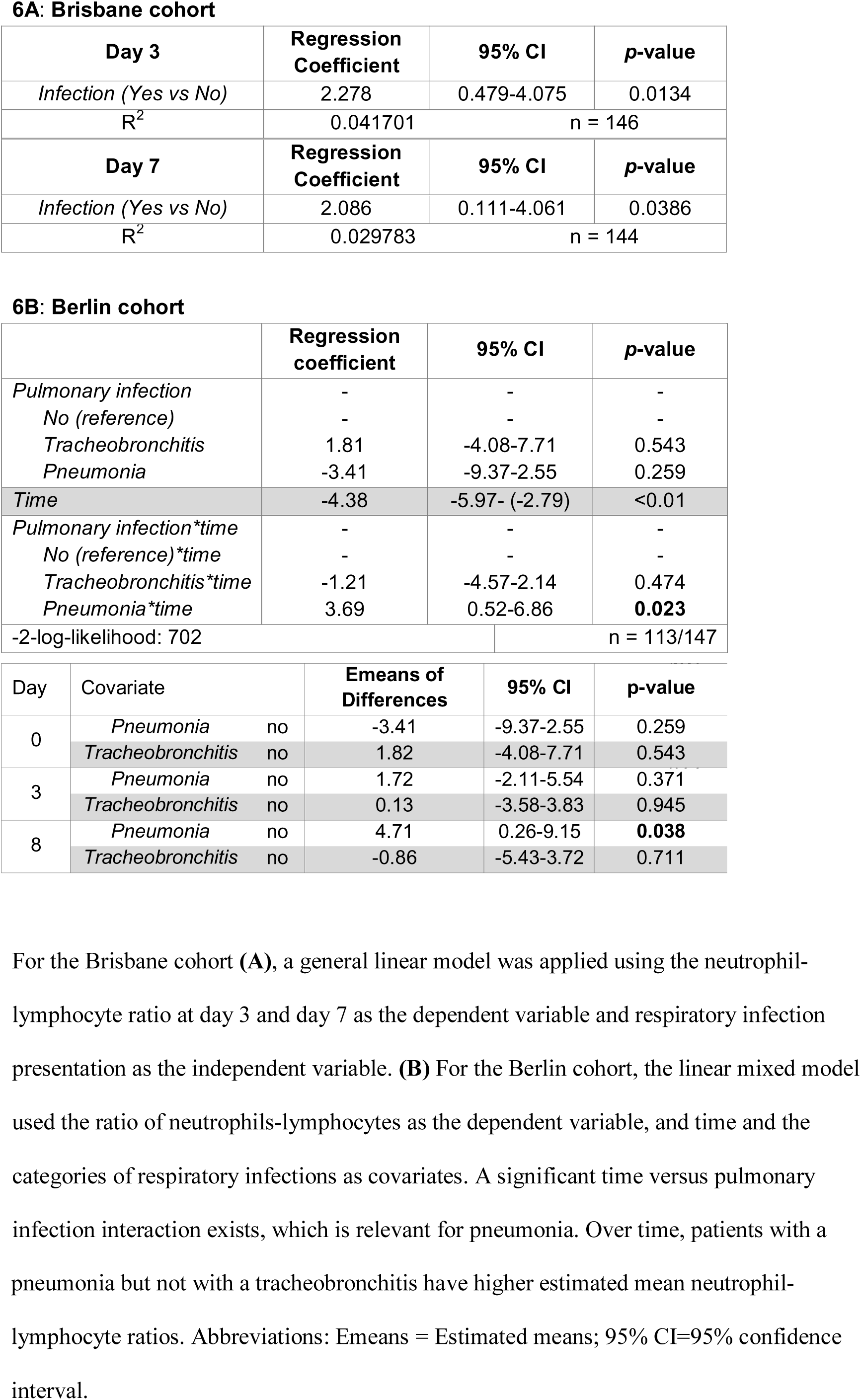
Association of neutrophil-lymphocyte ratios and respiratory infection after SCI

| Day 3 | Regression Coefficient | 95% CI | p-value |
| --- | --- | --- | --- |
| *Infection (Yes vs No)* | 2.278 | 0.479-4.075 | 0.0134 |
| R<sup>2</sup> | 0.041701 | n = 146 |  |
| Day 7 | Regression Coefficient | 95% CI | p-value |
| *Infection (Yes vs No)* | 2.086 | 0.111-4.061 | 0.0386 |
| R<sup>2</sup> | 0.029783 | n = 144 |  |

**6B: Berlin cohort**
|  | Regression coefficient | 95% CI | p-value |
| --- | --- | --- | --- |
| *Pulmonary infection* | - | - | - |
| *No (reference)* | - | - | - |
| *Tracheobronchitis* | 1.81 | -4.08-7.71 | 0.543 |
| *Pneumonia* | -3.41 | -9.37-2.55 | 0.259 |
| *Time* | -4.38 | -5.97- (-2.79) | <0.01 |
| *Pulmonary infection\*time* | - | - | - |
| *No (reference)\*time* | - | - | - |
| *Tracheobronchitis\*time* | -1.21 | -4.57-2.14 | 0.474 |
| *Pneumonia\*time* | 3.69 | 0.52-6.86 | **0.023** |
| -2-log-likelihood: 702 |  |  | n = 113/147 |

| Day | Covariate | Emeans of Differences | 95% CI | p-value |
| --- | --- | --- | --- | --- |
| 0 | *Pneumonia* no | -3.41 | -9.37-2.55 | 0.259 |
|  | *Tracheobronchitis* no | 1.82 | -4.08-7.71 | 0.543 |
| 3 | *Pneumonia* no | 1.72 | -2.11-5.54 | 0.371 |
|  | *Tracheobronchitis* no | 0.13 | -3.58-3.83 | 0.945 |
| 8 | *Pneumonia* no | 4.71 | 0.26-9.15 | **0.038** |
|  | *Tracheobronchitis* no | -0.86 | -5.43-3.72 | 0.711 |

For the Brisbane cohort (**A**), a general linear model was applied using the neutrophil-lymphocyte ratio at day 3 and day 7 as the dependent variable and respiratory infection presentation as the independent variable. (**B**) For the Berlin cohort, the linear mixed model used the ratio of neutrophils-lymphocytes as the dependent variable, and time and the categories of respiratory infections as covariates. A significant time versus pulmonary infection interaction exists, which is relevant for pneumonia. Over time, patients with a
pneumonia but not with a tracheobronchitis have higher estimated mean neutrophil-lymphocyte ratios. Abbreviations: Emeans = Estimated means; 95% CI=95% confidence interval.

Although neutrophil numbers alone were not indicative and/or predictive of respiratory infections during the acute phase of SCI, a positive relationship between the neutrophil-lymphocyte ratio and infection presentation was evident in both the Brisbane and Berlin patient cohorts. In the Brisbane cohort, neutrophil-lymphocyte ratio were typically higher at 3 dpi in patients that were either experiencing or developing such an adverse event (p=0.0373; *Figure 5G*). A general linear model further confirmed that infection incidence is linked to the neutrophil-lymphocyte ratio, not only at 3 but also 7 dpi (3dpi: R^2^ (95% CI) = 0.042 (0.48 – 4.08), p=0.0134; 7dpi: R^2^ (95% CI) = 0.030 (0.11 – 4.06), p=0.0386; *Table 6A*). This finding was recapitulated in the Berlin cohort, where a linear mixed model also showed a significant association between an elevated neutrophil-lymphocyte ratio and an increased incidence of pneumonia at 7 dpi (*Table 6B, Figure 5N*). These findings suggest that a WBC profile with neutrophilia and a low lymphocyte count either increases respiratory infection risk, or is an indicator of infection presentation/development. To explore whether such a WBC profile may dictate and/or reflect the severity of infection, we assessed the length of stay for individual patients with respiratory infections in the intensive care unit (ICU) as a proxy. In doing so, we found that the neutrophil-lymphocyte ratio at 3 dpi was positively correlated with the length of stay in ICU (r^2^ = 0.1593, p=0.0073; *Figure 5H*); a similar albeit weaker association was found for the neutrophil-monocyte ratio at this timepoint (r^2^ = 0.0958, p=0.0409; *Figure 5H*). To better illustrate the temporal relationship between WBC changes and infection presentation, we also superimposed the documented day of onset onto the neutrophil-lymphocyte ratios for the first week post-injury, which showed that the vast majority of patients acquired such an adverse event 3-4 days after hospitalisation (*Supplementary e-Figure 5*). Taken together, we found no evidence within the Brisbane cohort for lymphopenia, trauma severity and/or initial AIS grading as being possible risk factors per se for the increased infection susceptibility in patients with a high-level SCI. However, we did uncover that a greater acute systemic neutrophil response is correlated with respiratory infection presentation in SCI patients, particularly when considered in conjunction with the lymphocyte count, and this finding was reproduced in both the Brisbane and Berlin patient cohorts.

## DISCUSSION

We conducted a retrospective study to document early WBC changes in patients with a traumatic SCI, and to uncover potential relationships with concomitant injury factors and neurological outcomes. Significant results from the derivation cohort were subsequently tested in an independent validation cohort to increase the robustness of our observations. We show that the neutrophil response to SCI is influenced by trauma severity and predictive of long-term neurological recovery. We further confirm previous observations that lymphocyte numbers are decreased in the first week following injury, a finding that has generally been interpreted as an indicator of systemic insufficiency in adaptive immunity. However, the phenomenon of lymphopenia itself was not found to be positively associated with the presentation of respiratory infections in high-level SCI patients. However, a heightened neutrophil-lymphocyte ratio did coincide with respiratory infection occurrence, suggesting that the combination of a greater acute neutrophil response and reduced lymphocytes may predispose to and/or signify the development of such adverse events after traumatic SCI.

### The acute myeloid response to SCI is hallmarked by systemic neutrophilia

The role of neutrophils in recovery from SCI has been extensively investigated in experimental animal models, although some controversy remains as to their precise contribution.^9,30,31^ We ourselves demonstrated that the extent to which neutrophils are mobilised and recruited to the lesion site is directly and inversely correlated with outcomes in SCI mice.^8^ These findings are in agreement with the general consensus that, without immunomodulation, the presence of these cells in the injured spinal cord promotes additional damage through production and release of cytotoxic factors including pro-inflammatory cytokines, proteases, peroxidase activity and reactive oxidative species.^32,33^ The present findings support the premise that an injurious role for neutrophils also extends to human patients. Specifically, the circulating neutrophil count acutely after SCI was highly associated with injury severity, lesion level, and the neurological outcome. This inverse relationship between circulating neutrophil numbers at presentation and the longer-term neurological outcome remained when corrected for NISS / initial AIS grading, sex and age and infection presentation (*Table 3 and Supplementary e-Table 4*).^8^ Subgroup analysis of the derivation (i.e. Brisbane) cohort revealed two distinct patient groups, i.e. those presenting with a pronounced acute neutrophilia after SCI compared to others in which the circulating neutrophil count was never elevated above the normal reference range during the first 7 dpi. Why these divergent neutrophil profiles exist and/or what factors might be producing these differences is presently unclear but, importantly, patients that did not display an acute SCI-associated neutrophilia were far more likely to convert to an improved AIS grade, and this effect was reciprocated in the validation cohort. Neutrophils thus remain an important therapeutic target for intervention in acute SCI and future studies should now investigate how their phenotype is changed by SCI compared to the normal physiological state to allow for immunomodulation.

Circulating monocyte numbers mostly stayed within the normal reference range, but they were increased at both 1 and 7 dpi relative to admission for the Brisbane cohort; they were also highest at 8 dpi in the Berlin cohort. It is presently unclear if differential mobilization and/or recruitment of diverging monocyte subsets contributes to this temporal profile. Our previous work in mice showed that pro-inflammatory (i.e. Ly6C^hi^) monocytes are preferentially recruited to sites of SCI during the first week of injury,^34^ but routine hospital blood tests do not resolve different monocyte subsets. A more detailed characterisation of the human WBC compartment after SCI is therefore required as part of prospective studies.^35^

### Lymphopenia during the acute phase of SCI may be neuroprotective

Consistent with previous human and animal descriptions of SCI-IDS,^12,15,17,36-38^ a depression in lymphocyte numbers was observed in both the Brisbane and Berlin patient cohorts, most prominently at 1 and/or 3 dpi. Experimental animal studies have demonstrated that this reduction in lymphocytes after SCI, including in secondary immune organs such as the spleen and lymph nodes, is caused by a neuro-endocrine dysregulation.^17^ Current dogma dictates that disruption of neural pathways regulating sympathetic outflow from the thoracic spinal cord in high-level SCI (above T6) drives and/or predisposes to lymphopenia.^17^ We did not observe such a dependency on lesion level within the derivation cohort, at least not during the more acute phase of injury; no relationship between trauma severity and the occurrence of lymphopenia was observed either. Interestingly, we did find that patients with an isolated cervical SCI and acute clinical lymphopenia in the Brisbane cohort were more likely to have a positive conversion in AIS grade between admission and discharge. These data indicate that SCI-IDS, if defined by a reduced lymphocyte count at the level of the blood, is not necessarily deleterious for patient outcomes. This outcome aligns with previous pre-clinical work demonstrating better outcomes in SCI mice lacking T and/or B cells.^39,40^ Due to study design constraints, we were not able to validate this finding in the Berlin patient cohort, but further interrogation of lymphocyte subsets and their role in influencing recovery from SCI is warranted.

### Infection presentation after SCI and its relationship to the systemic WBC response

Consistent with previous literature,^29,41^ patients with cervical SCIs were found to be more at risk of respiratory infections than those with a thoracic lesion. Intubation was also positively linked to infection prevalence in both the Brisbane and Berlin patient cohorts, which is consistent with the fact that mechanical ventilation impedes effective airway secretion clearance.^42^ However, both this and paralysis of key muscles involved in airway clearance and breathing cannot fully explain the increased infection susceptibility of SCI patients.^17,18,43^ Indeed, SCI-IDS is considered a major contributor to this phenomenon and reductions in the circulating lymphocyte count are often used as an indicator or proxy for such immune insufficiency. Yet, we found no link between lymphopenia and infection presentation in our patient cohorts. This complements recent mouse work which provided evidence that it is not lymphopenia itself but rather a disruption of noradrenalin and cortisol homestasis due to adrenal gland dysfunction that drives the increased incidence of pneumonia in high-level SCI.^17^ We also did not find any evidence in this study for impaired neurological recovery in infected patients, at least not on the basis of a positive conversion in AIS grade, nor was there an association with having a complete SCI (i.e. AIS grade A). These findings appear somewhat at odds with previous reports where acquired infections were predictive of increased mortality and/or poorer long-term motor recovery.^14-16,18^ In addition to the inherent limitations that come with a smaller sample size (*Supplementary e-Table 4C*), the use of more stringent and/or sensitive measures of motor and sensory control in these prior studies is likely to have revealed more subtle differences in recovery that may go undetected with AIS grade conversion analysis. Likewise, there is disparity in the accepted definition of pneumonia in the clinical setting that can render the comparison of infection presentation and outcomes between studies unreliable.^44^ Because of the traumatic nature of SCI, a chest radiograph is often insufficient without an extended review of other clinical and microbiological signs to formulate a complete diagnosis.^45,46^ A more standardised approach for evaluating acquired infections and comparing these prospectively between independent centres is therefore required.

Lastly, we did find that the incidence of pneumonia in cervical SCI patients was higher in those that presented with clinical neutrophilia on admission. Neutrophil-lymphocyte ratios were also significantly higher at 3dpi in Brisbane patients that developed respiratory infections. An increase in the neutrophil-lymphocyte ratio for SCI patients developing pneumonia was also detected in the Berlin cohort. This correlative link between neutrophils numbers and infection presentation may be explained by the well-known involvement of neutrophils in combating infections. On the other hand, it is also known that systemic inflammatory response syndrome (SIRS) can cause tissue damage in distant organs such as the lungs.^30^ It is therefore tempting to speculate that abnormal neutrophil activation and/or dysfunction following high-level SCI may create a microenvironment that allows for infections to take hold. A caveat is, however, the fact that infections themselves typically also induce neutrophilia, making it difficult to determine cause and effect. Plotting the neutrophil-lymphocyte ratio against the documented day of infection onset did, however, not only reveal an immediate surge in respiratory infections 1 day after SCI, i.e. when neutrophil-lymphocyte ratios are high, but also a second wave when neutrophil numbers had mostly receded again to within range. Whether the magnitude of the neutrophil mobilization, SIRS and/or SCI-IDS effects on neutrophils are contributing factors to this phenomenon remains to be elucidated, and experimental animal studies could explore this more directly.

### Concluding remarks

We demonstrate prognostic value for early neutrophil and lymphocyte counts as long-term predictors for patient outcomes, specifically when used in conjuction with concomitant injury factors such as trauma severity, injury level, age and sex. Our study design did not include a critically ill patient groups without neurological injury and hence we cannot comment on the specificity of the observed WBC responses for SCI. Regardless, the associations between WBC numbers and patient outcomes still stand. With a pressing need to better manage and treat acute SCI, novel multi-factorial analysis models that can stratify and more accurately predict patient outcomes as presented here and by others^47^ have great value, not just for assessing the efficacy of new interventions, but also to identify targets for future therapies.

## ETHICAL APPROVAL

The Brisbane cohort dataset was approved by the Human Research Ethics Committee for Metro South Health (HREC/16/QPAH/196) and ratified by The University of Queensland (HREC #2014000031). The Berlin cohort dataset (COaT-SCI and observational studies) were locally approved by the Human Research Ethics Committee of Charité-Universitätsmedizin Berlin (#EA2/015/15 and #EA1/001/09, respectively).

## ABBREVIATIONS

AIS: American Spinal Injury Association Impairment Scale
ANOVA: analysis of variance
ASIA: American Spinal Injury Association
AUC: area under the curve
BMI: body mass index
C1-8: cervical vertebra 1-8
CDC: Centres for Disease Control and Prevention
COaT-SCI: Comparative Outcome And Treatment Evaluation in Spinal Cord Injury
dpi: days post-injury
ICU: intensive care unit
IQR: interquartile range
ISNCSCI: International Standards for Neurological Classification of Spinal Cord injury
MT: multi-trauma
NISS: new injury severity score
OR: odds ratio
PAH: Princess Alexandra Hospital
ROC: receiver operator characteristic
SCI: spinal cord injury
SCI-IDS: spinal cord injury-induced immune depression syndrome
SIRS: systemic inflammatory response syndrome
T1-12: thoracic vertebra 1-12
WBC: white blood cell

## AUTHOR CONTRIBUTIONS

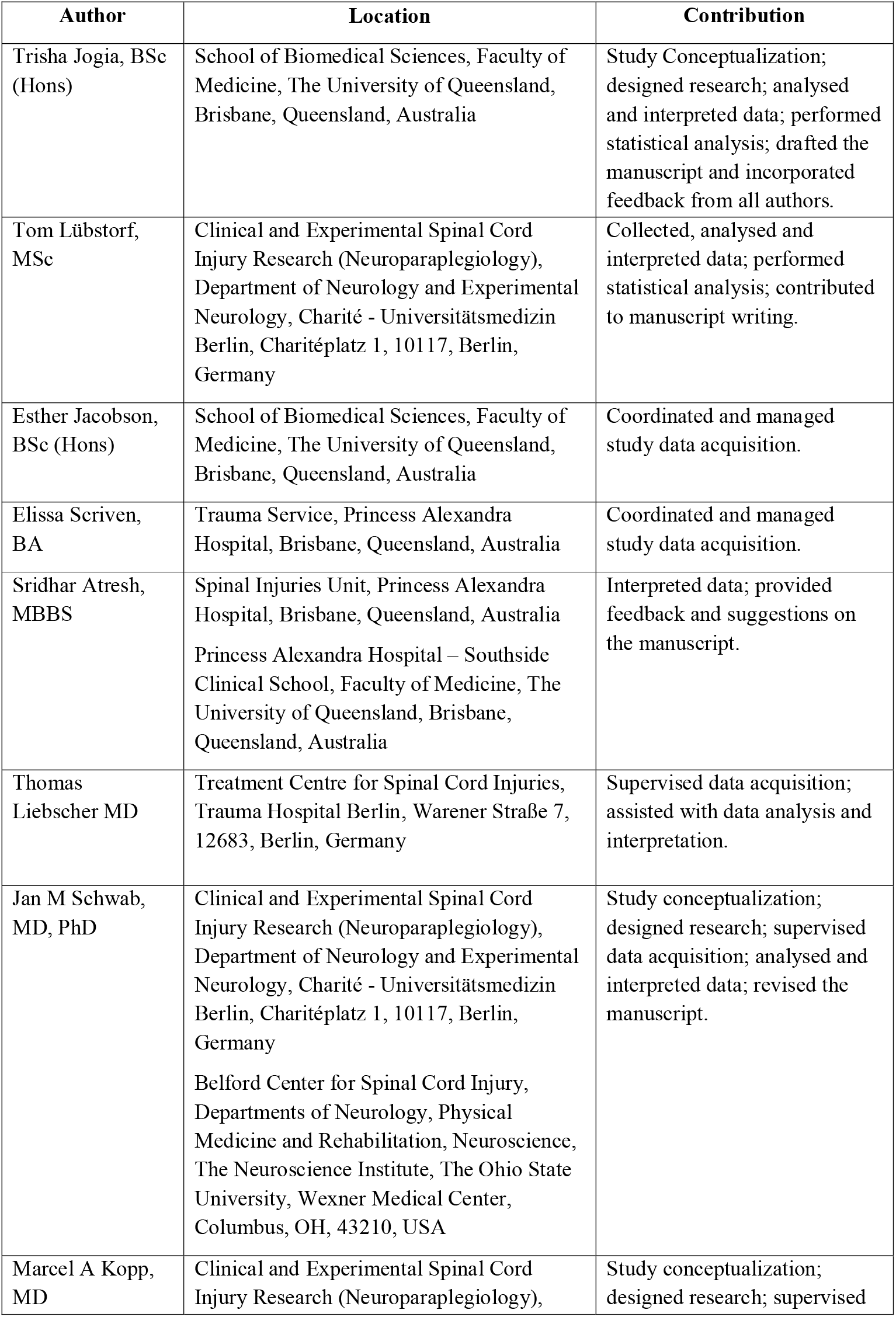
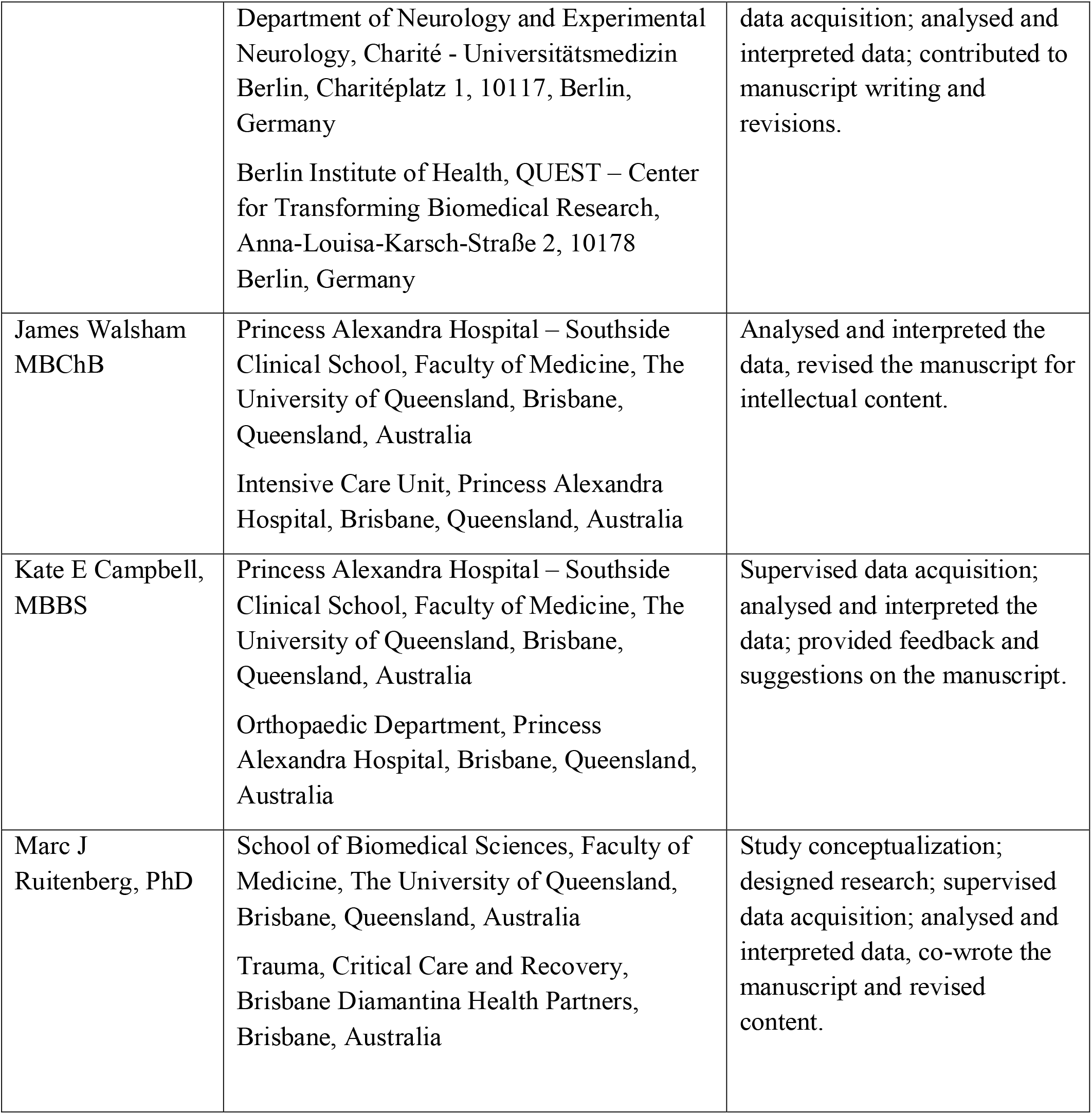

---

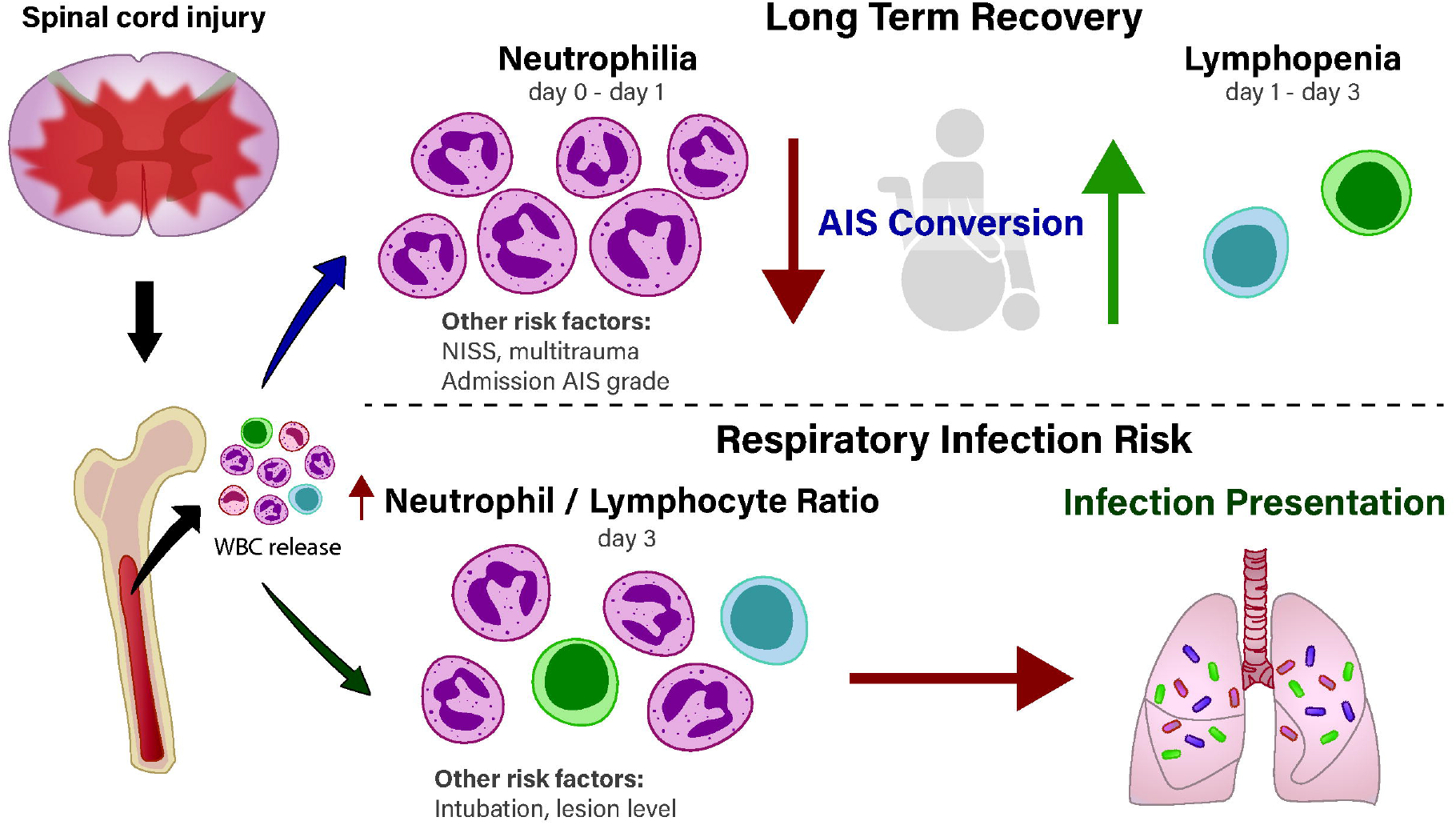

