## Supplementary Material for "Prognostic value of early leukocyte fluctuations for recovery from traumatic spinal cord injury"

### Supplementary Tables

**Supplementary e-Table 1:** patient characteristics and analysis details in the Berlin cohort

**A: Timing of blood collection after SCI**

| | Days after SCI, median (IQR) |
|---|---|
| *Visit 1, n=33* | 0 (0-1) |
| *Visit 2, n=43* | 3 (2-4) |
| *Visit 3, n=48* | 8 (7-9) |

**B: Screening for a possible selection bias.**

| | n | Bivariate models | | Multiple model | |
|---|---|---|---|---|---|
| Covariates | | OR (95% CI) | p-value | OR (95% CI) | p-value |
| *Age, per one year increase* | 324 | 1.01 (0.99-1.03) | 0.139 | 1.01 (0.99-1.03) | 0.306 |
| *BMI, per one point increase* | 314 | 0.99 (0.92-1.05) | 0.673 | 0.98 (0.91-1.05) | 0.482 |
| *Sex, male* | 324 | 0.46 (0.2-1.07) | 0.072 | 0.55 (0.23-1.32) | 0.182 |
| *AIS, grade A* | 318 | 1.69 (0.92-3.13) | 0.092 | 1.67 (0.82-3.41) | 0.161 |
| *Neurological level* | 316 | - | 0.711 | - | 0.949 |
| *lumbosacral* | - | 0.79 (0.38-1.63) | 0.52 | 0.98 (0.4-2.37) | 0.960 |
| *thoracic* | - | 1.14 (0.5-2.57) | 0.759 | 1.14 (0.49-2.64) | 0.766 |
| *cervical (reference)* | - | - | - | - | - |
| *Accompanying injury, yes* | 324 | 0.78 (0.42-1.45) | 0.431 | 0.79 (0.39-1.6) | 0.510 |
| Nagelkerkes $R^2$=0.044 | | | | | n=306 |

**(A)** Blood collection timepoints for the Berlin cohort. **(B)** A logistic regression model was calculated to assess the risk of selection bias due to an uneven distribution of demographic and clinical characteristics between eligible and excluded patients as dependent variable (eligible = 0, excluded = 1) and with age, BMI, AIS, neurological level, sex and accompanying injury as covariates. Abbreviations: American Spinal Cord Injury Association Impairment Scale; BMI = body mass index; IQR = interquartile range; OR = odds ratio; SCI = spinal cord injury; 95% CI=95% confidence interval.

**Supplementary e-Table 2*:*** Association of neutrophil count at day 3 with AIS-conversion in the Berlin cohort

**A: All patients**

| | **Unadjusted OR** | **95% CI** | ***p*-value** |
|---|---|---|---|
| *Neutrophil count at day 3* | 1.5 | 1.09-2.07 | **0.013** |
| Nagelkerke's $R^2$ | 0.281 | | n = 39 |
| | **Adjusted OR** | **95% CI** | ***p*-value** |
| *Neutrophil count at day 3* | 1.63 | 1.08-2.45 | **0.02** |
| *Age* | 1.08 | 1.00-1.15 | **0.029** |
| *Sex, male* | 0.38 | 0.01-11.1 | 0.574 |
| *AIS, grade A* | 0.54 | 0.08-3.59 | 0.525 |
| *Level* | | | |
| *Lumbosacral* | 1.02 | 0.06-16.77 | 0.989 |
| *Thoracic* | 4.57 | 0.47-44.46 | 0.190 |
| *Cervical (reference)* | - | - | - |
| *Multi-trauma, yes* | 0.58 | 0.07-4.67 | 0.615 |
| *BMI* | 0.81 | 0.58-1.13 | 0.211 |
| Nagelkerke's $R^2$ | 0.385 | | n = 38 |

**B: Excluding patients with admission AIS grade D**

| | **Unadjusted OR** | **95% CI** | ***p*-value** |
|---|---|---|---|
| *Neutrophil count at day 3* | 2.11 | 1.22-3.65 | **0.008** |
| Nagelkerke's $R^2$ | 0.552 | | n = 28 |

Logistic regression models using AIS grade conversion as the dependent variable (improvement = 0, non-improvement =1) and neutrophil count at day 3 post-injury as the primary predictor variable. **(A)** Unadjusted univariate model with continuous frequency of neutrophils as the covariate, and multivariate model adjusted with age, BMI, AIS grade, neurological level, sex and accompanying injury (multi-trauma) as covariates. **(B)** Univariate model with continuous neutrophil counts as covariate for patients with an admission AIS grade of A, B and C only. Abbreviations: AIS = American Spinal Cord Injury Association Impairment Scale; BMI = body mass index; OR = odds ratio; 95% CI = 95% confidence interval.

**Supplementary e-Table 3*:*** Association of neutrophilia with AIS grade conversion in the Berlin-cohort including patients AIS D

**All patients**

| All patients | **Unadjusted OR** | **95% CI** | ***p*-value** |
|---|---|---|---|
| *Neutrophilia, yes* | 6.00 | 1.12-32.28 | **0.037** |
| Nagelkerke's $R^2$ | 0.172 | | n = 40 |
| | **Adjusted OR** | **95% CI** | ***p*-value** |
| *Neutrophilia, yes* | 10.58 | 1.03-108.46 | **0.047** |
| *Age* | 1.07 | 1.01-1.14 | **0.022** |
| *Sex, male* | 0.41 | 0.01-12.58 | 0.607 |
| *AIS, grade A* | 0.63 | 0.11-3.61 | 0.605 |
| *Level* | | | |
| *Lumbosacral* | 1.53 | 0.11-22.05 | 0.754 |
| *Thoracic* | 2.81 | 0.36-22.16 | 0.328 |
| *Cervical (reference)* | - | - | - |
| *Multi-trauma, yes* | 0.60 | 0.09-4.08 | 0.597 |
| *BMI* | 0.89 | 0.68-1.16 | 0.387 |
| Nagelkerke's $R^2$ | 0.385 | | n = 39 |

Neutrophilia was defined as the blood cell count being above the normal range. AIS grade conversion was used as the dependent variable (improvement = 0, non-improvement = 1) and neutrophilia as the primary predictor variable in a univariate model. The multivariate model was adjusted for age, sex, admission AIS grade, neurological level of the lesion, associated multi-trauma and BMI. For AIS A, approximately 15-20% AIS grade conversions can be expected. For incomplete SCI, the expected conversion rate for AIS B is 50-70%, 80-95% for C, and 2-40% for D (Fawcett et al. 2007). As part of a sensitivity analysis, all incomplete SCIs (i.e. admission AIS grades B, C and D) were combined here; results without inclusion of AIS grade D patients is presented in Table 3B of the main document. Abbreviations: AIS = American Spinal Cord Injury Association Impairment Scale; BMI = body mass index; OR = odds ratio; 95% CI=95% confidence interval.

**Supplementary e-Table 4:** Association of neutrophil count differences and the neurological outcome from SCI

**4A: Day 0 – day 1 difference**

| | **Unadjusted OR** | **95% CI** | ***p*-value** |
|---|---|---|---|
| *Day 0 – day 1 difference* | 0.934 | 0.865-1.009 | 0.0847 |
| Nagelkerke's $R^2$ | 0.0475 | | n=93 |
| | **Adjusted OR** | **95% CI** | ***p*-value** |
| *Day 0 – day 1 difference* | 1.078 | 0.900-1.291 | 0.4167 |
| *Admission neutrophil count* | 0.803 | 0.671-0.961 | **0.0166** |
| *Age* | 0.999 | 0.936-1.036 | 0.9574 |
| *Sex, female* | 0.293 | 0.054-1.579 | 0.1532 |
| *Admission AIS grade* | | | **0.0016** |
| *AIS A* | 0.066 | 0.015-0.298 | **0.0125** |
| *AIS B* | 0.127 | 0.021-0.766 | 0.3761 |
| *AIS C (reference)* | **-** | - | - |
| *Level* | | | 0.2948 |
| *Cervical* | 0.821 | 0.169-3.972 | 0.1516 |
| *Lumbosacral* | 16.746 | 0.379-739.862 | 0.1227 |
| *Thoracic (reference)* | - | - | - |
| *Multi-trauma, yes* | 0.423 | 0.130-1.378 | 0.1535 |
| *Respiratory infection* | 1.445 | 0.419-4.983 | 0.5597 |
| Nagelkerke's $R^2$ | 0.4256 | | n=93 |

**4B: Day 1 – day 3 difference**

| | **Unadjusted OR** | **95% CI** | ***p*-value** |
|---|---|---|---|
| *Day 1 – day 3 difference* | 0.864 | 0.759-0.984 | **0.0278** |
| Nagelkerke's $R^2$ | 0.0803 | | n=93 |
| | **Adjusted OR** | **95% CI** | ***p*-value** |
| *Day 1 – day 3 difference* | 0.765 | 0.635-0.921 | **0.0047** |
| *Admission neutrophil count* | 0.831 | 0.723-0.955 | **0.0089** |
| *Age* | 0.992 | 0.955-1.031 | 0.6993 |
| *Sex, female* | 0.321 | 0.055-1.876 | 0.2071 |
| *Admission AIS grade* | | | **0.0037** |
| *AIS A* | 0.067 | 0.013-0.331 | **0.0144** |
| *AIS B* | 0.132 | 0.021-0.832 | **0.4031** |
| *AIS C (reference)* | **-** | - | - |
| *Level* | | | 0.1847 |
| *Cervical* | 0.981 | 0.185-5.196 | 0.0978 |
| *Lumbosacral* | 124.342 | 0.679-999.999 | 0.0661 |
| *Thoracic (reference)* | - | - | - |
| *Multi-trauma, yes* | 0.393 | 0.635-0.921 | 0.1600 |
| *Respiratory infection* | 1.287 | 0.341-4.858 | 0.7099 |
| Nagelkerke's $R^2$ | 0.5014 | | n=91 |

**4C: Day 3 – day 7 difference**

| | Unadjusted OR | 95% CI | *p*-value |
|---|---|---|---|
| *Day 3 – day 7 difference* | 1.344 | 1.117-1.618 | **0.0018** |
| Nagelkerke's $R^2$ | 0.1590 | | n=99 |
| | **Adjusted OR** | **95% CI** | ***p*-value** |
| *Day 3 – day 7 difference* | 1.556 | 1.166-2.077 | **0.0027** |
| *Admission neutrophil count* | 0.842 | 0.738-0.959 | **0.0098** |
| *Age* | 1.010 | 0.976-1.046 | 0.5724 |
| *Sex, female* | 0.145 | 0.019-1.108 | 0.0627 |
| *Admission AIS grade* | | | **0.0003** |
| *AIS A* | 0.017 | 0.002-0.127 | **0.0016** |
| *AIS B* | 0.047 | 0.006-0.394 | 0.2363 |
| *AIS C (reference)* | **-** | - | - |
| *Level* | | | 0.0650 |
| *Cervical* | 0.273 | 0.048-1.537 | **0.0194** |
| *Lumbosacral* | 23.706 | 0.585-960.086 | **0.0447** |
| *Thoracic (reference)* | - | - | - |
| *Multi-trauma, yes* | 0.442 | 0.124-1.580 | 0.2093 |
| *Respiratory infection* | 3.501 | 0.776-15.796 | 0.1031 |
| Nagelkerke's $R^2$ | 0.5724 | | n=97 |

Logistic regression models used AIS grade conversion as the dependent variable (non-improvement = 0, improvement =1) and the difference in neutrophil count between the two specified timepoints (neutrophilia resolution) as the primary predictor variable. **(A)** Difference between day 0 (admission) and day 1 neutrophil counts, **(B)** difference between day 1 and day 3 neutrophil counts, and **(C)** the difference between day 3 and day 7 neutrophil counts. Multivariate models were adjusted for the admission neutrophil count, age, sex, admission AIS grade, neurological level of the lesion, associated multi-trauma and respiratory infection presentation. Abbreviations: AIS = American Spinal Cord Injury Association Impairment Scale; BMI = body mass index; OR = odds ratio; 95% CI=95% confidence interval.

**Supplementary e-Table 5:** Association between lymphopenia and neurological recovery in the Berlin cohort

**A: All patients**

| | **Unadjusted OR** | **95% CI** | ***p*-value** |
|---|---|---|---|
| *Lymphopenia, yes* | 1.57 | 0.41-5.96 | 0.504 |
| Nagelkerke's $R^2$ | 0.015 | | n=40 |
| | **Adjusted OR** | **95% CI** | ***p*-value** |
| *Lymphopenia* | 1.56 | 0.31-7.75 | 0.589 |
| *Age* | 1.05 | 1.0-1.1 | 0.064 |
| *BMI* | 0.93 | 0.73-1.18 | 0.543 |
| *AIS A* | 0.36 | 0.07-1.92 | 0.232 |
| *Level* | | | 0.724 |
| *Thoracic* | 2.04 | 0.39-14.14 | 0.472 |
| *Lumbosacral* | 0.97 | 0.11-8.91 | 0.979 |
| *Cervical (reference)* | - | - | - |
| *Sex, male* | 1.05 | 0.05-23.55 | 0.977 |
| *Multitrauma, yes* | 0.45 | 0.08-2.62 | 0.376 |
| Nagelkerke's $R^2$ | 0.299 | | n = 39 |

**B: Excluding patients with admission AIS grade D**

| | **Unadjusted OR** | **95% CI** | ***p*-value** |
|---|---|---|---|
| *Lymphopenia, yes* | 0.56 | 0.15-2.12 | 0.395 |
| Nagelkerke's $R^2$ | 0.035 | | n = 28 |

Logistic regression modelling was applied for the entire patient cohort without missing data using AIS conversion (non-conversion 0, conversion = 1). Lymphopenia was defined as a cell count at visit 1 or 2 below the normal response. **(A)** Univariate model with Lymphopenia as covariate. Multivariate model with Lymphopenia, age, BMI, AIS, neurological level, sex and accompanying injury as covariates. **(B)** Univariate model with Lymphopenia as covariate and Patients AIS A, B, and C patients only. Abbreviations: AIS=American Spinal Cord Injury Association impairment scale, BMI= body mass index, 95% CI=95% confidence interval

#### Supplementary Figures

##### Supplementary e-Figure 1: Diagram for the Berlin validation cohort.

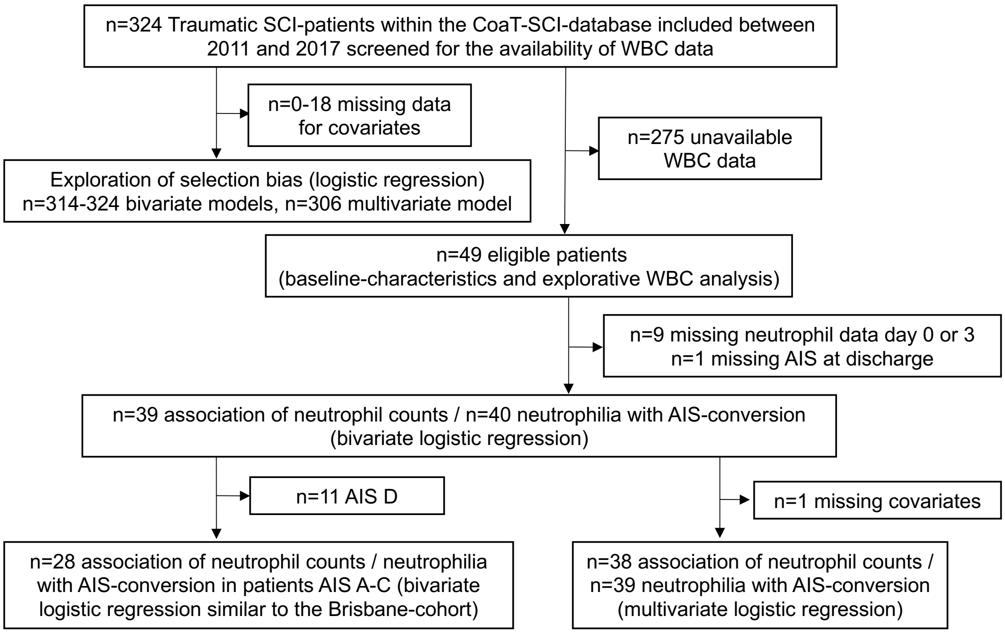


Patients with a traumatic spinal cord injury (SCI) and enrolled into the Comparative Outcome and Treatment Evaluation in SCI (COaT-SCI) study were screened for the availability of WBC data. These data were then tested for selection bias (see *Table 2*). Datasets used for specific statistical analyses as indicated depended on the subgroup being tested, sample size limitations and availability of data. Abbreviations: AIS = American Spinal Cord Injury Association Impairment Scale; WBC = white blood cell.

**Supplementary e-Figure 2: Influence of lesion level on conversion rates and the link between neutrophilia and monocyte counts.**

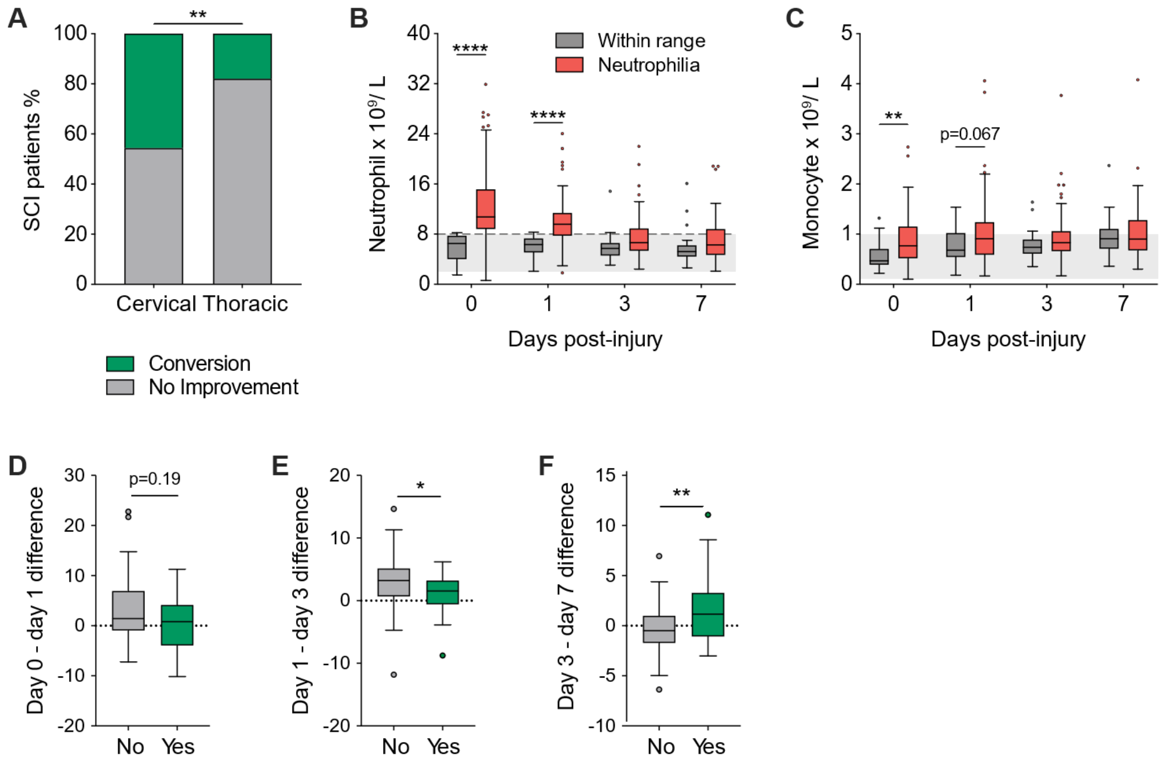


**(A)** AIS Grade conversion rates in patients with a cervical versus thoracic SCI (Fisher's exact test). **(B)** Temporal profile of circulating neutrophil numbers in SCI patients in those with and without neutrophilia (dashed grey line indicates the upper margin of the clinical reference range shown in light grey; two-way ANOVA with Bonferroni's *post hoc* test). **(C)** Circulating monocyte numbers in patients with and without acute neutrophilia (two-way ANOVA with Bonferroni's *post hoc* test). **(D-F)** Differences in neutrophil count between day 0 (admission) and day 1 **(D)**, day 1 and day 3 **(E),** and day 3 and day 7 **(F)** post-injury for patients with AIS grade conversion (Yes) and patients who showed no improvement (No; Mann-Whitney Test). Box and whisker plots show the median and interquartile range, Tukey whiskers and outliers indicated as dots. **p<0.01, ****p<0.0001.

**Supplementary e-Figure 3: Lymphopenia is not associated with trauma severity or the lesion level of SCI.**

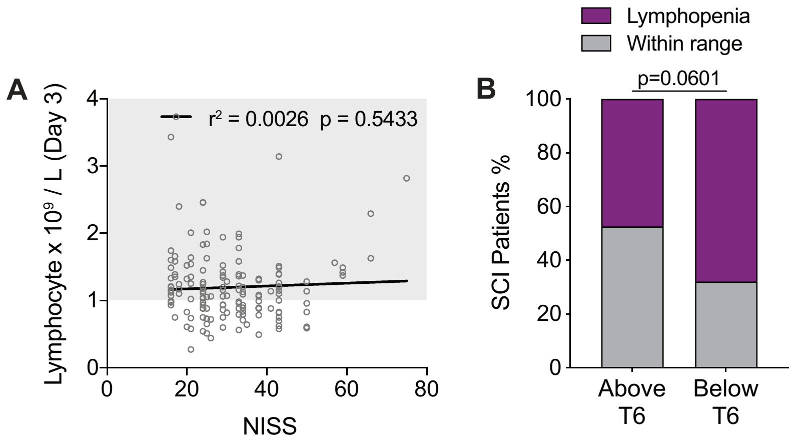


**(A)** NISS data indicative of trauma severity was not associated with circulating lymphocyte numbers at 3 dpi ($r^2$ value = Pearson's correlation coefficient, linear fit indicated by solid black line). **(B)** Incidence of lymphopenia in SCI patients with lesions above or below the level of T6 (Fisher's exact test). NISS = new injury severity score. T6=thoracic vertebra 6.

**Supplementary e-Figure 4: Sputum culture results, WBC and NISS data for SCI patients with and without acute respiratory infection presentation.**

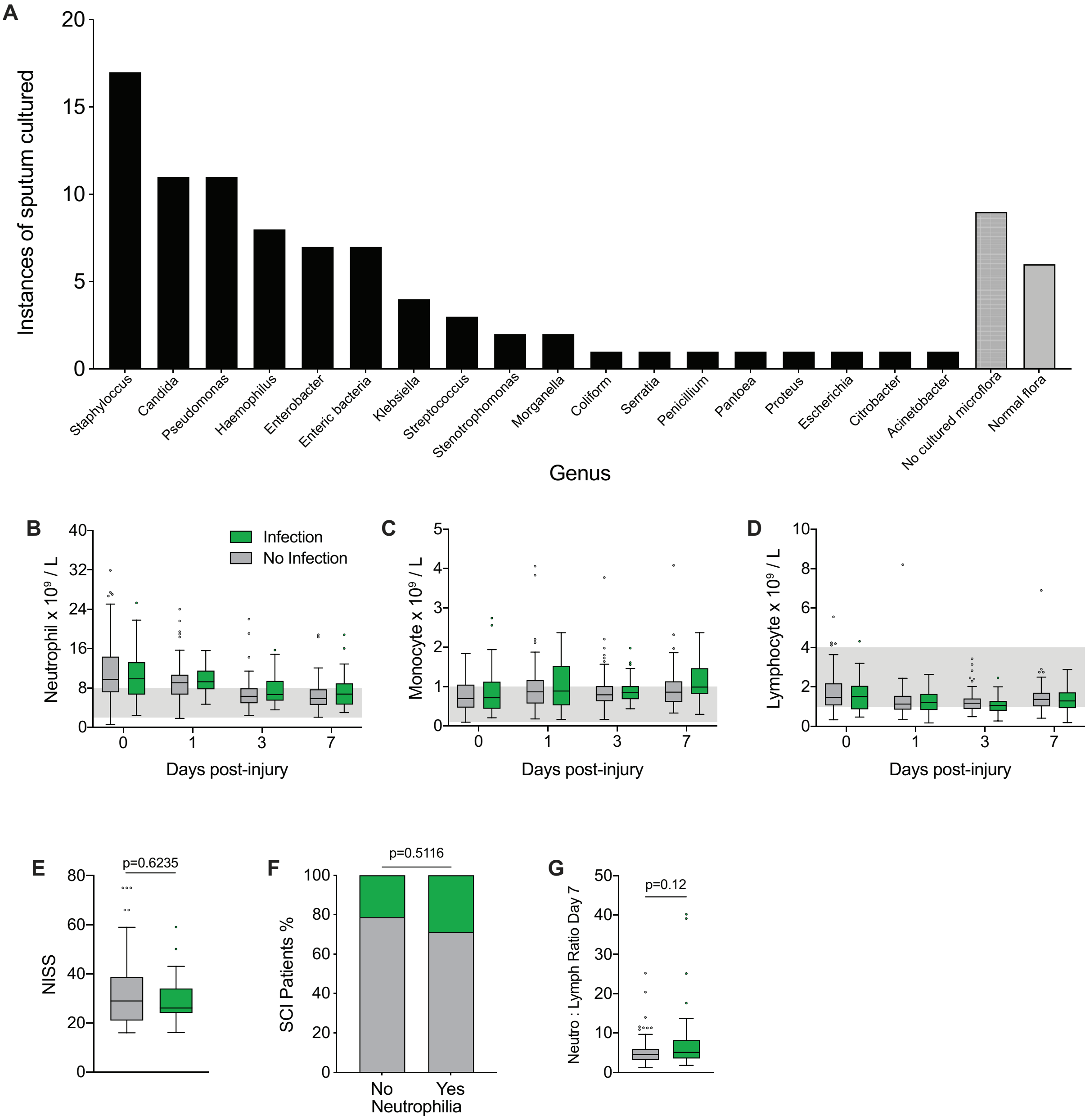


**(A)** 18 genera of microorganisms were cultured from the sputum of patients who developed an airway infection early after injury. The most common genus cultured was Staphylococcus, followed by Candida and Pseudomonas. **(B-D)** Circulating WBC numbers for infected versus non-infected patients during the first week after SCI (2-way ANOVA; ns). **(E)** NISS data for patients with and without airway infection presentation (Mann-Whitney Test). **(F)** Incidence of airway infection presentation for all Brisbane patients (i.e. regardless of lesion level) with and without neutrophilia (Fisher's exact test). **(G)** Neutrophil-lymphocyte ratios at 7 dpi for patients with and without respiratory infection. Box and whisker plots are shown with the median and interquartile range, Tukey whiskers, and with outliers indicated as dots. AIS = American Spinal Cord Injury Association Impairment Scale; dpi = days post-injury; NISS = new injury severity score; ns = not significant; WBC = white blood cell.

**Supplementary e-Figure 5: Temporal relationship between neutrophil-lymphocyte ratios and respiratory infection incidence / onset in the first week post-SCI.**

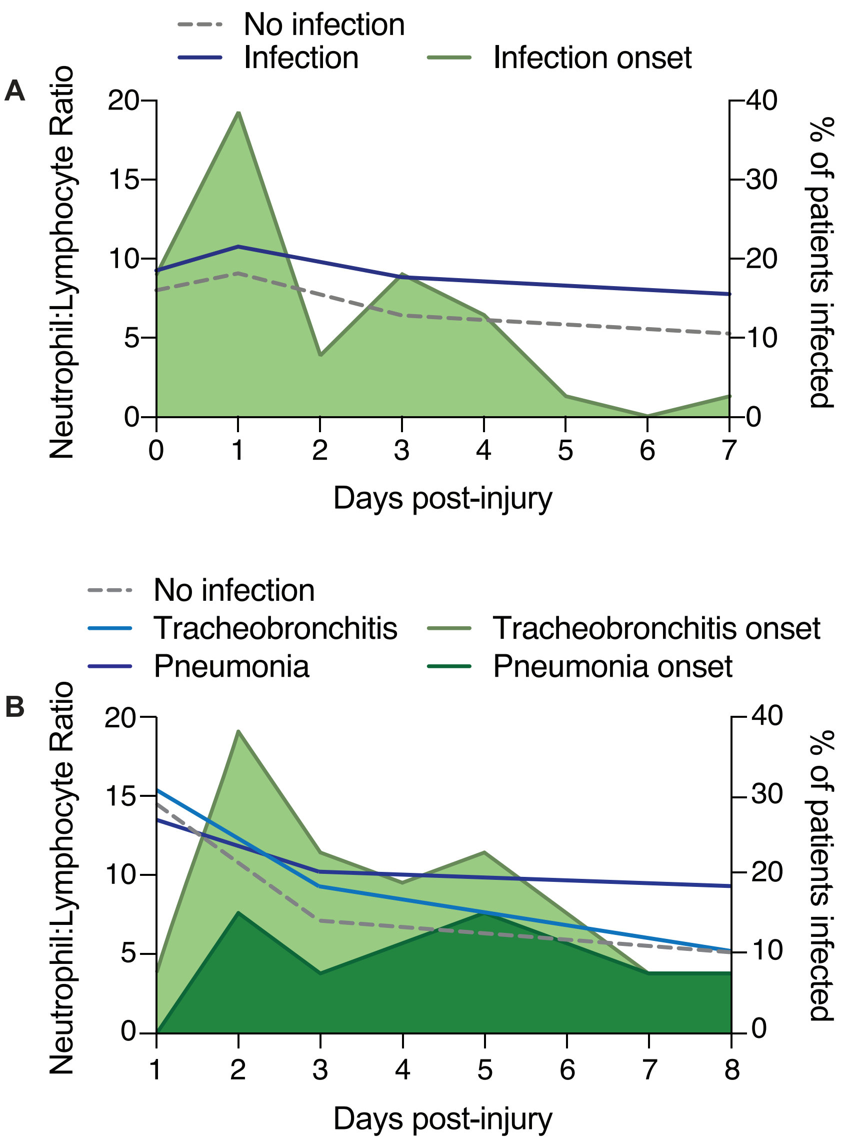


**(A)** Neutrophil-lymphocyte ratios for infected SCI patients in the Brisbane cohort (*left y-axis*; blue line) and documented time (dpi) of infection presentation (*right y-axis*; green). Neutrophil-lymphocyte ratios for SCI patients without a respiratory infection are indicated by the grey dashed line for comparison. **(B)** Matching data on neutrophil-lymphocyte ratios and the timing of infection presentation in the Berlin cohort presenting both patients with pneumonia (dark blue/green) and tracheobronchitis (light blue/green). Abbreviations: dpi = days post-injury; SCI = spinal cord injury.
